# Dynamic Quantum Clustering of Glioma RNA-seq Reveals Interpretable Tumor States Linked to Diagnosis and Survival

**DOI:** 10.64898/2026.04.09.26350535

**Authors:** Fereshteh Jahanbani, Rajan Maynard, Steven J. Schrodi, Marvin Weinstein

## Abstract

Public RNA-seq datasets can refine tumor diagnosis and outcome prediction, but biological heterogeneity, variable biopsy composition, and analytic drift often obscure clinically meaningful structure. We applied Dynamic Quantum Clustering (DQC), an unsupervised, geometry-preserving method requiring no clinical labels, to 692 TCGA gliomas. Whole-transcriptome DQC separated low-grade glioma– and glioblastoma-enriched states, while a DQC-derived 90-gene panel resolved three pure low-grade glioma subclusters with distinct survival outcomes and one glioblastoma-rich cluster with 97.1% positive predictive value. Four non-overlapping gene modules emerging from these clusters were used to define four biological coordinates (“BioCoords”) for each tumor. In BioCoord space, tumors formed a continuous clinical and molecular glioma trajectory, including a mesenchymal axis associated with progressively worse survival. The framework also preserved histologic and molecular relationships while exposing intermediate tumor states not captured by discrete labels. Projection of an independent, heterogeneous CGGA cohort of 693 gliomas into the same BioCoord space reproduced the principal low-dimensional organization, cluster relationships, and survival-associated continuum observed in TCGA. These findings support robust cross-cohort molecular stratification, improve classification across clinically and molecularly diverse gliomas, and provide a practical framework for prognosis, patient selection, and investigation of glioma progression.

**Highlights:**

- DQC reveals intrinsic RNA-defined organization across TCGA gliomas without using clinical labels.
- A 90-gene set resolves three LGG risk groups and one GBM-enriched group.
- Survival measures form distinct structures across mesenchymal and developmental Biocoord states.
- Biocoord space preserves diagnostic organization in a validation cohort despite incomplete gene overlap

## Introduction

Diffuse gliomas, ranging from lower-grade glioma (LGG) to glioblastoma (GBM) [1–4], exemplify the profound challenges of pursuing precision oncology to biologically heterogeneous tumors. Current adult diffuse glioma classification integrates histology with molecular markers such as IDH mutation and 1p/19q codeletion [5].

GBM is particularly difficult to manage because of diffuse brain infiltration, marked intratumoral heterogeneity, limited resectability, resistance to chemoradiation, and poor drug penetration across the blood–brain barrier [6–11]. Most tumors recur, and median survival remains approximately 15 months in contemporary treatment series [12] despite combined radiotherapy and temozolomide-based therapy [13]. Clinical decisions therefore depend on accurate diagnosis, early risk assessment, and the ability to recognize when a tumor may be moving toward a more aggressive state [14]. In particular, detecting when an LGG is acquiring GBM-like, treatment-resistant biology could influence the timing and intensity of surgery, radiation, systemic treatment, and clinical monitoring [5,15,16].

Histology, imaging, and targeted DNA-based molecular testing remain essential for glioma diagnosis and treatment planning. However, these tools also have practical limitations. A biopsy samples only part of a spatially heterogeneous tumor, and standard diagnostic categories may not fully capture the biological direction in which an individual tumor is moving. Tumors that share the same histologic or molecular label can still differ in growth behavior, treatment resistance, and clinical outcome [17]. RNA-based molecular profiling offers a complementary view because it captures active cellular programs, including proliferation, invasion, immune signaling, and lineage-state changes. Thus, RNA expression may help place tumors along biologically meaningful disease-state maps rather than describing them only as fixed diagnostic categories [18].

Despite the growing availability of large cancer transcriptomic resources [19–21], converting RNA-seq data into clinically useful maps of tumor progression remains difficult, converting RNA-seq data into clinically useful maps of tumor progression remains difficult. Datasets such as TCGA have shown that molecular features can define clinically meaningful glioma subgroups [15], while independent glioma cohorts such as CGGA provide an opportunity to test whether such structures are reproducible across patient populations, platforms, and preprocessing pipelines [22]. However, several obstacles limit this goal. First, gliomas are spatially heterogeneous, and site-limited sampling may obscure progression-related biology [23–25]. Second, diagnostic labels can vary across observers, institutions, and datasets, introducing noise into supervised analyses [26]. Third, technical differences, including batch effects, platform differences, tumor purity, and stromal or immune admixture, can cause computational maps to reflect dataset-specific artifacts rather than reproducible tumor biology [27–30]. Consequently, reproducing findings across independent datasets is intrinsically challenging, as cohort-specific noise obscures the shared biological structure [16,30]. Common unsupervised approaches such as PCA and t-SNE are useful for visualizing complex RNA-seq data [31,32]. However, they may emphasize the strongest sources of variation, even when those signals are not the most relevant to progression or outcome, and they may not preserve gradual biological relationships between tumor states [33–35]. For glioma, the key unmet need is therefore not simply another clustering method, but a way to convert RNA expression into an interpretable map that preserves both recognizable tumor groups and intermediate progression-related states.

To address this need, we developed DBUG, a DQC-based BioCoord unsupervised workflow centered on Dynamic Quantum Clustering (DQC), an unsupervised, geometry-preserving approach for resolving structure within complex, high-dimensional datasets [36–38]. DQC allows tumors with similar RNA-expression profiles [39] to organize into shared high-density regions and organize together before clinical or pathologic labels are introduced for interpretation [35,36]. This design is well suited to glioma because clinically recognizable categories, such as LGG and GBM, may coexist with gradual transitions in tumor biology, including changes in invasion, extracellular-matrix remodeling, developmental programs, and cellular state [38,39]. Rather than forcing every tumor into a predefined group, this framework first lets tumors organize by RNA similarity and then asks whether the resulting structure matches diagnosis, molecular markers, and patient outcome. This approach is intended to reduce sensitivity to cohort-specific noise while preserving tumor-state patterns that are reproducible and clinically meaningful [40].

We applied this workflow to bulk RNA-seq data from 692 TCGA gliomas [19] to ask whether RNA expression alone could reveal a clinically meaningful organization of glioma states. Clinical labels, molecular annotations, and survival data were added only after the RNA-defined structure was generated. Starting from the full transcriptome, the analysis identified a broad LGG-to-GBM-like organization, which was then refined into a compact 90-gene signal and translated into four biologically interpretable RNA-based coordinates, termed BioCoords. These coordinates were designed to place each tumor within a disease-state map rather than assign it only to a fixed cluster [41,42].

We then tested whether the TCGA-derived BioCoord structure could be recovered in an independent CGGA cohort with different gene coverage, preprocessing, and clinical composition. For this validation, the CGGA analysis was restricted at the outset to tumors with available overall-survival data, while survival and clinical annotations were still used only after RNA-based BioCoord calculation and clustering. Across TCGA and CGGA, BioCoord-defined tumor organization aligned with histology, molecular markers, and survival, supporting that the framework captures reproducible glioma biology rather than a cohort-specific pattern.

This framework is intended to complement, rather than replace, conventional histologic and molecular classification. Histology, IDH status, 1p/19q codeletion, MGMT promoter methylation, imaging, age, extent of resection, and treatment history remain essential for clinical decision-making. Instead of representing glioma only as fixed diagnostic categories, BioCoords place individual tumors within an interpretable progression-associated landscape that may help identify intermediate or discordant tumors that are difficult to classify by standard approaches. More broadly, this study provides a general strategy for extracting reproducible and clinically relevant biological structure from heterogeneous transcriptomic and other multi-omics datasets [43], with potential applications across cancer types and other molecularly complex diseases.

## Methods

### Translational Objective and Pipeline development Overview

The DBUG workflow was developed to convert bulk glioma RNA-seq data into an interpretable tumor-state map that preserves both discrete tumor groups and continuous progression-related structure. The goal was not to build a supervised classifier from predefined diagnostic or survival labels, but to determine whether RNA expression alone could reveal a biologically and clinically meaningful organization of glioma samples.

The workflow consisted of three sequential stages. First, DQC was applied in a fully unsupervised, label-free manner to map the broad RNA-based organization of gliomas across the LGG-to-GBM spectrum. Second, genes contributing little to the dominant tumor-state structure were filtered to reduce background variation and identify a compact 90-gene progression-associated panel. Third, the 90-gene structure was translated into four biological coordinates, termed BioCoords, which summarize coordinated RNA-expression programs and allow each tumor to be positioned within a reduced tumor-state space for clinical interpretation. Clinical variables, including histology, molecular markers, progression-free survival, and overall survival, were not used to generate the DQC maps, select the 90-gene panel, or calculate BioCoords. These annotations were added only after RNA-based structure generation to evaluate whether the resulting tumor organization aligned with clinically meaningful features. Because DBUG is an unsupervised structure-discovery framework rather than a supervised prediction model, we did not use clinical endpoints to train a classifier or optimize a train/test split, thereby avoiding issues that can arise in supervised modeling workflows, including data leakage, unrepresentative sampling, and overfitting [44–46]. Instead, reproducibility was assessed by applying the TCGA-derived BioCoord framework to an independent CGGA validation cohort with different gene coverage and preprocessing.

### Cohort and preprocessing

We analyzed TCGA glioma bulk RNA-seq data comprising 692 tumors with matched clinical metadata, including histology and time-to-event variables for progression-free survival and overall survival. All clustering, dimensionality reduction, and feature-selection steps were performed without using clinical labels or survival outcomes.

Expression matrices were restricted to 20,057 protein-coding genes, log₂-scaled, and L2-normalized per sample. L2 normalization was used to place samples on a comparable scale before distance-based clustering, reducing the influence of sample-level differences in overall expression magnitude.

For visualization, DQC frames were displayed using the first three principal components. These three components were used only to display the evolving tumor map and were not used as the sole input for clustering. Histology and survival annotations from TCGA were overlaid strictly post hoc after each unsupervised DQC run to assess diagnostic concordance and relate RNA-defined structures to clinical outcomes.

#### 1. DQC hyperparameters and visualization

DQC was used to organize tumors according to RNA-expression similarity without assigning a predefined number of clusters. Conceptually, the method allows tumors with similar expression profiles to move toward shared high-density regions, forming groups that represent related tumor states. This process preserves both discrete endpoint groups and intermediate structures that may reflect gradual biological transitions. An intuitive description of the DQC dynamics and illustrative models is provided in **Supplemental Note 1, DQC_Evolution_Dynamics** and **Figures S1–S3**.

For high-dimensional inputs, DQC used an internal dimensionality-reduction step to improve computational stability and reduce background variation. When the input contained more than 16 dimensions, including the full transcriptome, the 554-gene panel, and the 90-gene panel, the input matrix was reduced to 16 principal components before DQC clustering. The 16-component setting was selected during initial optimization to retain broad expression structure while reducing noise. For the four-dimensional BioCoord representation, no PCA reduction was applied; DQC was performed directly on the four BioCoord values.

To maintain consistency across analyses, the DQC mass parameter was held constant. The sigma parameter was scaled according to the variance of the input space at each stage to avoid collapse of the tumor structure [47]. DQC was applied sequentially to four feature representations: the full 20,057 protein-coding gene dataset, the unsupervised 554-gene panel, rank-ordered subsets ending in the 90-gene panel, and the four-dimensional BioCoord representation.

For each analysis, representative DQC frames were saved at the initial, intermediate, and final stages. Initial and intermediate frames were used to visualize how tumors organized into continuous structures, whereas final frames were used to define stable endpoint groups. All plotted DQC frames were displayed in PC1–PC3 for visualization only. The clustering itself was performed in the designated analysis space: 16 PCs for high-dimensional gene-expression inputs and four coordinates for the BioCoord representation. Robustness was assessed by repeating analyses with increased PCA dimensionality; the major tumor partitions were qualitatively preserved, indicating that the observed structures were not driven by the display settings.

Clinical variables, including histology, molecular markers, PFS, and OS, were withheld during DQC clustering and feature selection. These annotations were added only after RNA-based structures were generated, allowing diagnostic and prognostic relevance to be evaluated without using clinical labels to build the maps.

#### 2. Unsupervised feature selection to isolate core disease-progression signals from the protein-coding transcriptome

The initial DQC analysis of the full 20,057-gene dataset separated tumors into two primary RNA-defined groups. Because many genes in the transcriptome are unlikely to contribute meaningfully to this dominant LGG-to-GBM-like structure, we next asked whether background transcriptional variation could be reduced while preserving the core tumor organization.

For each gene, we calculated the absolute difference in mean normalized expression, |Δμ|, between the two DQC-derived groups. These groups were defined without clinical labels and were only later found, after post hoc annotation, to correspond to LGG-enriched and GBM-enriched tumor states. Genes were then ranked from smallest to largest |Δμ| to generate a continuous expression-difference curve. To identify a data-driven cutoff separating low-contribution genes from higher-signal genes, we compared the visual inflection point of this curve with its numerically computed first derivative. The cutoff was selected at the elbow of the curve, where the expression-difference values began to rise more sharply. On the log₂-scaled, L2- normalized data, this threshold occurred at |Δμ| ≈ 0.0036, retaining 554 genes from the original 20,057 protein-coding genes.

To evaluate whether this focused gene set preserved the tumor structure, we compressed the 554-gene expression profiles for all 692 tumors into 16 principal components and re-ran DQC using the same mass parameter and proportionately scaled sigma parameter described above. TCGA histology and time-to-event variables were then added strictly post hoc. Concordance metrics, including overall accuracy and positive predictive value, were calculated to evaluate alignment between the 554-gene DQC groups and clinical LGG/GBM diagnoses. Kaplan–Meier estimators and log-rank tests were used to evaluate progression-free survival and overall survival across the RNA-defined groups.

To interpret the biology associated with the two 554-gene DQC groups, we performed differential expression analysis on the normalized expression data using Welch’s t-test with Benjamini–Hochberg correction across all tested genes. Genes with FDR < 0.05 were considered significant. Volcano plots were used to display expression differences between groups and corresponding statistical significance. Canonical pathway enrichment was then assessed using Ingenuity Pathway Analysis with Benjamini–Hochberg-adjusted statistics [48,49]. These differential-expression and pathway analyses were performed only after clustering to interpret the resulting tumor states; they were not used for DQC clustering, feature selection, or threshold determination.

#### 3. Dimensionality reduction to identify a compact 90-gene progression-associated panel

Bulk RNA-seq captures thousands of genes, many of which may reflect baseline cellular functions, sample composition, or background variation rather than the dominant tumor-state structure. We therefore tested whether further reducing the 554-gene panel could preserve the clinically relevant glioma organization while producing a smaller and more interpretable gene set.

Genes from the 554-gene panel were ranked using Welch’s t-test comparing the two primary DQC groups identified after the elbow-based filtering step. The resulting p-values were used only to rank genes for feature ordering; they were not used as hypothesis tests or to define statistical significance at this stage. This ranking approach is conceptually similar to using moderated differential-expression statistics, such as the B statistic, to prioritize genes for downstream analysis [50].

To evaluate progressively smaller gene sets, we assembled top-ranked panels of 140, 120, 100, and 90 genes and re-ran DQC on each reduced expression matrix using the same mass parameter, proportionately scaled sigma parameter, and 16-component input representation described above. Across the evaluated panels, DQC consistently preserved the broad LGG–GBM separation, while revealing three final groups with the 140- and 120-gene panels and four groups with the 100- and 90-gene panels.

The 90-gene panel was selected as the final compact feature set because it preserved clear separation into four tumor groups while reducing the feature set to the smallest tested panel that maintained stable cluster structure. Further reduction below 90 genes reduced cluster stability and weakened the separation between groups. The four DQC-defined groups from the 90-gene panel were designated clusters A, B, C, and D for downstream analysis. Clinical interpretation was performed only after clustering. TCGA histology was overlaid post hoc to assess diagnostic composition and cluster purity, and Kaplan–Meier analyses with two-sided log-rank tests were used to evaluate progression-free survival and overall survival across the four RNA-defined groups.

#### 4. Translating transcriptomic structure into biological coordinates: construction of BioCoords

After identifying the four-cluster structure from the 90-gene panel, we next asked whether this RNA-defined organization could be summarized by a smaller number of interpretable biological programs. To do this, we performed selected pairwise comparisons among the four DQC-defined clusters to identify non-overlapping gene modules, which were then used to construct four biological coordinates, termed BioCoords.

Based on the RNA-defined cluster organization and its post hoc clinical interpretation, we selected four comparisons that captured the major A→D→C→B tumor-state pattern with limited redundancy: A–B and D–B, representing broader LGG-to-GBM-like differences, and A–C and C–D, representing differences among intermediate LGG-enriched states. The remaining comparisons, A–D and B–C, were not used because they added limited independent structure. For each selected comparison, all 20,057 protein-coding genes were scored using a two-sample t-test between the two DQC-defined clusters. The resulting p-values were used only to rank genes for module construction; they were not used as formal hypothesis tests or for statistical significance at this step.

To keep each coordinate compact while retaining enough genes to represent a biological program, we selected the top 30 genes from each ranked comparison. Overlapping genes were assigned to the comparison in which they ranked highest, producing four non-overlapping modules containing 24 genes for A–B, 25 genes for A–C, 27 genes for D–B, and 19 genes for C–D, for a total of 95 BioCoord genes.

For each module, the corresponding gene-expression values were extracted from the full TCGA dataset. Each gene was standardized across all tumors by z-scoring, calculated by subtracting the cohort mean and dividing by the cohort standard deviation. This placed genes on a common scale and prevented genes with larger expression ranges from dominating the module score. Genes with zero variance would have been excluded; none were present. The standardized expression values were then averaged within each module to generate one coordinate value per tumor. Combining the four module scores produced a four-dimensional BioCoord profile for each of the 692 TCGA tumors. This approach is conceptually related to gene-set scoring and pathway-based representations, but the BioCoord modules were derived from the RNA-defined tumor structure rather than from predefined pathway labels or clinical categories [42,51–54].

DQC was then applied directly to the four-dimensional BioCoord matrix to test whether the tumor organization observed with the 90-gene panel was preserved in this reduced biological-coordinate representation. Because the BioCoord input contained only four variables, the 16-PC reduction step used for higher-dimensional gene-expression matrices was not applied. DQC was therefore performed directly on the four BioCoord values for each tumor. The mass parameter was held constant, and sigma was scaled according to the input variance as described above.

All DQC runs, feature rankings, and BioCoord calculations were performed without using histology, molecular annotations, PFS, or OS labels. Clinical variables were added only after BioCoord construction and clustering to evaluate diagnostic concordance and survival associations. Because each BioCoord was defined by an explicit, non-overlapping gene module, this representation allowed tumor position to be interpreted in relation to specific biological programs, including mesenchymal/extracellular-matrix remodeling and developmental gene-expression patterns.

For clinical evaluation, TCGA histology was overlaid after BioCoord-based clustering to compute concordance metrics. Associations between BioCoord-derived groups and clinical outcomes were evaluated using Kaplan–Meier analyses for progression-free survival and overall survival, measured in days from diagnosis to event or last follow-up, with two-sided log-rank tests. Samples with missing time-to-event data were excluded from the corresponding survival analysis.

#### 5. Cross-sample set validation in the CGGA glioma dataset

To test whether the TCGA-derived tumor organization could be recovered in an independent cohort, we analyzed glioma RNA-seq data from the Chinese Glioma Genome Atlas (CGGA). The full CGGA cohort included 693 tumors comprising lower-grade gliomas and glioblastomas, including both primary and recurrent tumors.

CGGA clinical and molecular metadata, including histology, tumor status, WHO grade, IDH mutation status, 1p/19q codeletion status, MGMT promoter methylation status, treatment variables, progression-free survival, and overall survival, were downloaded from the CGGA website and are provided in **Table S6**.

Because the cross-cohort validation included post hoc survival assessment, survival analyses were restricted to CGGA tumors with available overall-survival data. Overall survival was not used to calculate BioCoords, perform clustering, or define tumor groups. Instead, OS and other clinical annotations were added only after RNA-based BioCoord calculation and DQC clustering to evaluate whether the resulting tumor organization aligned with clinical outcome.

Consistent with the TCGA analysis, all CGGA clustering and coordinate-construction steps were performed without using clinical labels, molecular annotations, treatment variables, or survival outcomes. These annotations were used only after the RNA-based structures had been generated.

CGGA gene-expression data were analyzed using the provided normalized expression matrix. Because TCGA and CGGA differed in gene coverage, sequencing pipelines, preprocessing, and annotation, gene-level validation was restricted to the 16,154 genes shared between the two cohorts. We did not explicitly correct for inter-cohort differences before validation; instead, these differences were treated as part of the real-world cross-cohort test. DQC was first applied to the 16,154 shared genes to assess whether direct shared-gene clustering could recover prognostic structure in CGGA. We then applied a knee-based denoising step that reduced the CGGA expression matrix to approximately 636 genes and repeated DQC using this reduced gene set. For gene-expression matrices, inputs were reduced to 16 principal components before DQC, consistent with the TCGA workflow.

Next, CGGA tumors were mapped into the four-dimensional BioCoord space defined from the TCGA analysis. Of the 95 TCGA-defined BioCoord genes, 78 were available in the CGGA dataset. For each CGGA tumor, BioCoord values were calculated from CGGA RNA expression by standardizing the available genes within each retained BioCoord module and averaging the standardized expression values within that module. This generated a four-coordinate BioCoord profile for each CGGA tumor using only CGGA RNA-expression data.

DQC was then applied directly to the four-dimensional CGGA BioCoord matrix. Because this representation contained only four variables, the 16-PC reduction step used for higher-dimensional gene-expression inputs was not applied. Intermediate DQC frames were used to visualize how tumors organized within BioCoord space, whereas final convergence frames defined stable BioCoord-derived groups for post hoc clinical evaluation.

After BioCoord calculation and DQC clustering were completed, the resulting BioCoord-derived cluster assignments were integrated with the CGGA clinical and molecular metadata. This integrated table, linking each CGGA tumor to its BioCoord- derived group and corresponding clinical annotations, is provided in **Table S7**. Data from **Table S7** was then used to interrogate the relationship between BioCoord- derived tumor groups and CGGA clinical features, including IDH mutation status, WHO grade, treatment context, and overall survival.

Survival differences between BioCoord-derived groups were evaluated using Kaplan– Meier estimators and two-sided log-rank tests, restricted to tumors with available OS data. Overall survival was also plotted directly within BioCoord space to evaluate whether tumor position in the RNA-based map tracked with clinical outcome. To determine whether BioCoord-derived groups reflected known glioma prognostic features, we examined the distribution of IDH mutation status, WHO grade, and OS across BioCoord clusters using the integrated annotations in **Table S7**.

To place the BioCoord findings in the context of standard clinical and molecular predictors, we also evaluated OS across established CGGA annotations, including WHO grade, IDH mutation status, 1p/19q codeletion status, MGMT promoter methylation status, age group, and treatment context. These analyses were used to assess whether conventional clinical or molecular variables explained survival heterogeneity and to determine whether BioCoord organization provided complementary RNA-based information beyond individual markers.

#### 6. General statistics, software, and reproducibility

Differential expression and canonical pathway enrichments (IPA) were evaluated with multiple-testing correction using the Benjamini–Hochberg procedure, defining significance at a false discovery rate (FDR) < 0.05. In contrast, the two-sample t-test p-values utilized in Sections 3 and 4 were employed strictly as monotonic metrics for feature ordering and module construction; no statistical inference or multiple-testing corrections were applied at those stages. DQC and the elbow-based gene selection were fully unsupervised and did not use histology or outcome labels. The inflection point for the elbow-based gene selection (|Δμ| ≈ 0.0036 on log₂-scaled, L2- normalized data) was determined primarily by a first-derivative scan to identify the point of maximal curvature; the automated Kneedle algorithm [55] was used solely as a secondary computational check and yielded a concordant threshold. Throughout the study, principal component analysis (PCA) served two distinct purposes: (1) an initial 16-component dimensionality reduction applied exclusively to large gene-expression matrices (>16 features) to optimize DQC computational efficiency, and (2) an independent 3-component rotation (PC1–PC3) utilized strictly for the 3D visualization of DQC evolutionary frames.

Analyses were performed using the Maple version of DQC [59] and standard Python/R environments (for statistics and plotting), utilizing IPA for canonical pathway enrichment. The exact preprocessing pipelines (log₂ scaling, L2 normalization), DQC hyperparameters (including the fixed mass and variance-scaled sigma parameters), random seeds, and custom scripts for feature ranking, module de-duplication, coordinate construction, and *post hoc* clinical evaluations are provided in the Extended Data/Code to ensure full reproducibility.

To summarize, cross-cohort validation was performed in the independent CGGA dataset using the same analytical framework. Analyses were restricted to overlapping genes between datasets (*n* = 16,154), and BioCoord projections were computed using available genes without imputation (78 of 95 retained). Feature selection procedures were applied independently within each cohort and used only for ordering or dimensionality reduction. DQC parameters (mass and variance-scaled sigma) and preprocessing steps were applied consistently across datasets. Survival analyses were performed independently within each cohort using Kaplan–Meier estimators. These steps ensured that reproducibility reflects the robustness of the underlying biological structure rather than the transfer of dataset-specific models.

Rather than transferring a supervised prediction model from TCGA to CGGA, the analysis tested whether a TCGA-derived biological coordinate framework could preserve clinically meaningful tumor organization in an independent cohort.

## Results

All clustering and gene-selection steps were performed without using histology, progression-free survival (PFS), or overall survival (OS) labels. Clinical metadata were added only after each analytical step to evaluate how well the RNA-derived tumor groups matched diagnosis and outcome.

### 1. A label-free DQC-BioCoords framework maps glioma progression from RNA expression alone

To identify the natural RNA-based organization of glioma, we designed a five-stage DQC-BioCoords workflow that progressively reduces transcriptomic complexity while preserving disease-relevant structure (**Figure 1**). The workflow begins with transcriptome-wide DQC analysis of 20,057 protein-coding genes to map the broad tumor organization without predefined labels. It then filters the data to isolate a 554- gene core signal, refines this signal into an optimal ∼90-gene panel, translates the resulting tumor structure into four biological coordinates (“BioCoords”), and finally tests whether the same structure can be recovered in an independent CGGA cohort. Each stage was designed to answer a specific biological question. The initial whole-transcriptome analysis asked whether gliomas contain an intrinsic RNA-based organization. The 554-gene filtering step asked whether this broad structure could be denoised to isolate a core LGG-to-GBM progression signal. The ∼90-gene refinement asked whether this signal could be compressed into a compact and clinically interpretable panel. The BioCoords step asked whether tumor groups could be translated into biological axes useful for prognosis and understanding the underlying molecular pathophysiology. Finally, independent cohort projection tested whether the resulting biological module map was reproducible.

**Figure 1.**
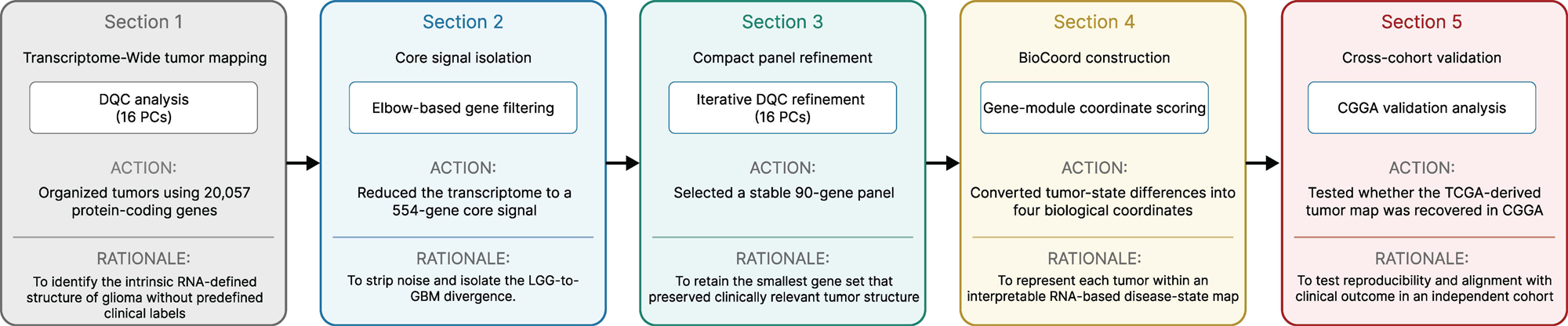
DBUG workflow for label-free mapping of glioma tumor states from RNA expression. Overview of the five-stage DBUG workflow used to convert glioma RNA-seq data into an interpretable tumor-state map. First, DQC was applied to 20,057 protein-coding genes to identify intrinsic RNA-defined glioma structure without predefined clinical labels. Second, elbow-based filtering reduced the transcriptome to a 554-gene core signal while preserving the LGG-to-GBM-like organization. Third, iterative DQC refinement identified a compact 90-gene panel that retained clinically relevant tumor structure. Fourth, tumor-state differences were translated into four biological coordinates, termed BioCoords, to represent each tumor within an interpretable RNA-based disease-state map. Fifth, the TCGA-derived BioCoord framework was tested in the independent CGGA cohort to evaluate reproducibility and clinical alignment. Histology, molecular annotations, treatment variables, and survival outcomes were withheld during clustering and feature selection and were added only afterward for clinical interpretation.

This roadmap therefore serves as the reference framework for the study results, linking each computational step to its biological purpose: moving from high-dimensional RNA-seq data toward a compact, interpretable, and externally testable map of glioma progression.

### 2. Whole-transcriptome DQC separates gliomas into LGG-enriched and GBM-enriched tumor states

We first applied DQC to RNA-seq data from 692 TCGA gliomas using the full transcriptome of 20,057 protein-coding genes after log₂ scaling and L2 normalization. This analysis asked whether RNA expression alone could reveal a natural organization of glioma samples.

For visualization, the DQC process was projected into the first three principal components. Even in the initial distribution, before final clustering, the tumors formed a broad cloud with two visible regional groups (**Figure 2A**). As DQC progressed, this broad distribution condensed into an extended, continuous one-dimensional trajectory (**Figure 2B**). At convergence, this trajectory resolved into two stable high-density tumor groups composed of 686 samples, along with six isolated tumor samples (**Figure 2C**).

**Figure 2.**
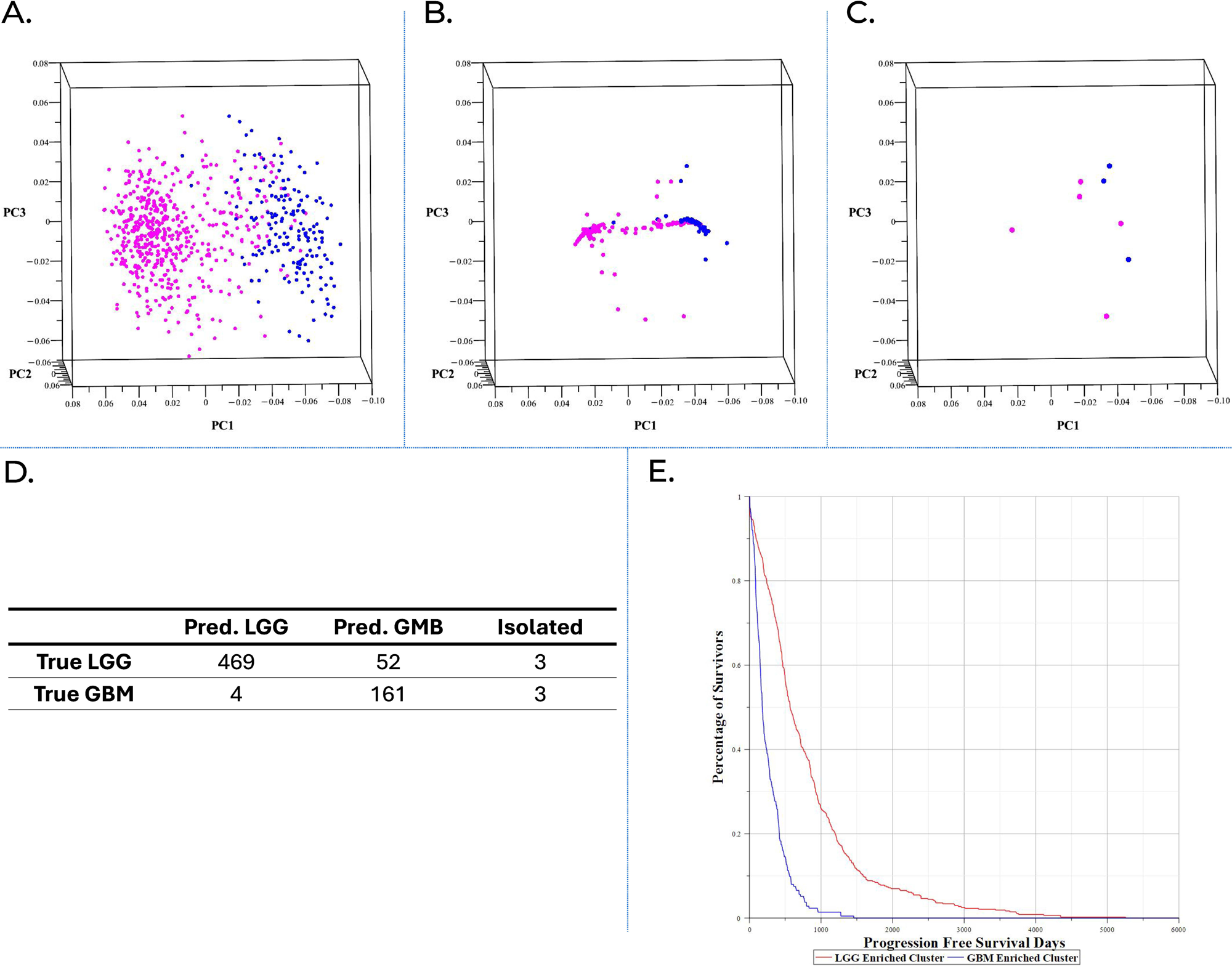
Transcriptome-wide DQC reveals two clinically meaningful glioma states from RNA expression alone. DQC was applied to RNA-seq data from 692 TCGA gliomas using all 20,057 protein-coding genes to test whether transcriptomic information alone could organize tumors in a way that was relevant to diagnosis and patient outcome. Histology, survival, and other clinical information were not used to define the groups. The first three principal components are shown only to visualize how the tumor map changes during DQC; each point represents the whole-transcriptome profile of one tumor, and tumors with more similar profiles appear closer together. (A) Starting DQC frame, in which tumors are organized into two broad regions based on their transcriptomic profiles. (B) Intermediate DQC frame, showing the tumors condensing into a continuous trajectory between the two emerging states. (C) Final DQC frame at convergence, showing two dense RNA-defined tumor groups and six isolated tumors. TCGA histology was added only afterward for interpretation: magenta, lower-grade glioma (LGG); blue, glioblastoma (GBM). (D) Post hoc comparison with TCGA histology, showing 91.8% concordance among the 686 tumors assigned to a main DQC group; six tumors remained isolated. (E) Kaplan–Meier analysis testing whether the RNA-defined groups also differed in progression-free survival. The GBM-enriched group had significantly shorter progression-free survival than the LGG-enriched group (two-sided log-rank P < 0.001). Together, these panels show how tumor patterns identified from RNA expression alone can be evaluated for their relevance to diagnosis and patient outcome.

When TCGA histology was subsequently added, the two DQC-defined groups showed strong diagnostic concordance. Cluster 1 was overwhelmingly enriched for LGG tumors, containing 469 of 473 LGGs, or 99.2%. Cluster 2 was enriched for GBM tumors, containing 161 of 213 GBMs, or 75.6%. Using histology only as an external benchmark, the label-free DQC assignment achieved an overall diagnostic accuracy of 90.9%, corresponding to 630 of 692 tumors (**Figure 2D**). Mean normalized expression values for all 20,057 genes across these initial DQC-predicted LGG- and GBM-enriched groups are provided in **Table S1**.

Survival analysis further supported the clinical relevance of this RNA-derived structure. Kaplan–Meier curves showed that patients in the LGG-enriched group had significantly longer PFS and OS than patients in the GBM-enriched group (P<1E-30) (**Figure 2E**). Together, these findings show that whole-transcriptome DQC can identify a clinically meaningful glioma organization without using clinical labels during clustering. This provided the foundation for reducing the gene set and isolating the core glioma progression-related signal.

### 3. Denoising the transcriptome reveals a core progression signal with stronger clinical separation

The whole-transcriptome analysis identified the broad structure of glioma, but it also included many genes that may reflect redundant, background, or noise-dominated variation. To focus on the genes most responsible for separating the two major tumor states, we filtered the transcriptome using the expression differences between the two primary DQC clusters as detailed in the **Methods**. This process identified a focused 554-gene glioma trajectory panel (**Figure 3A**, **Table S2**).

**Figure 3.**
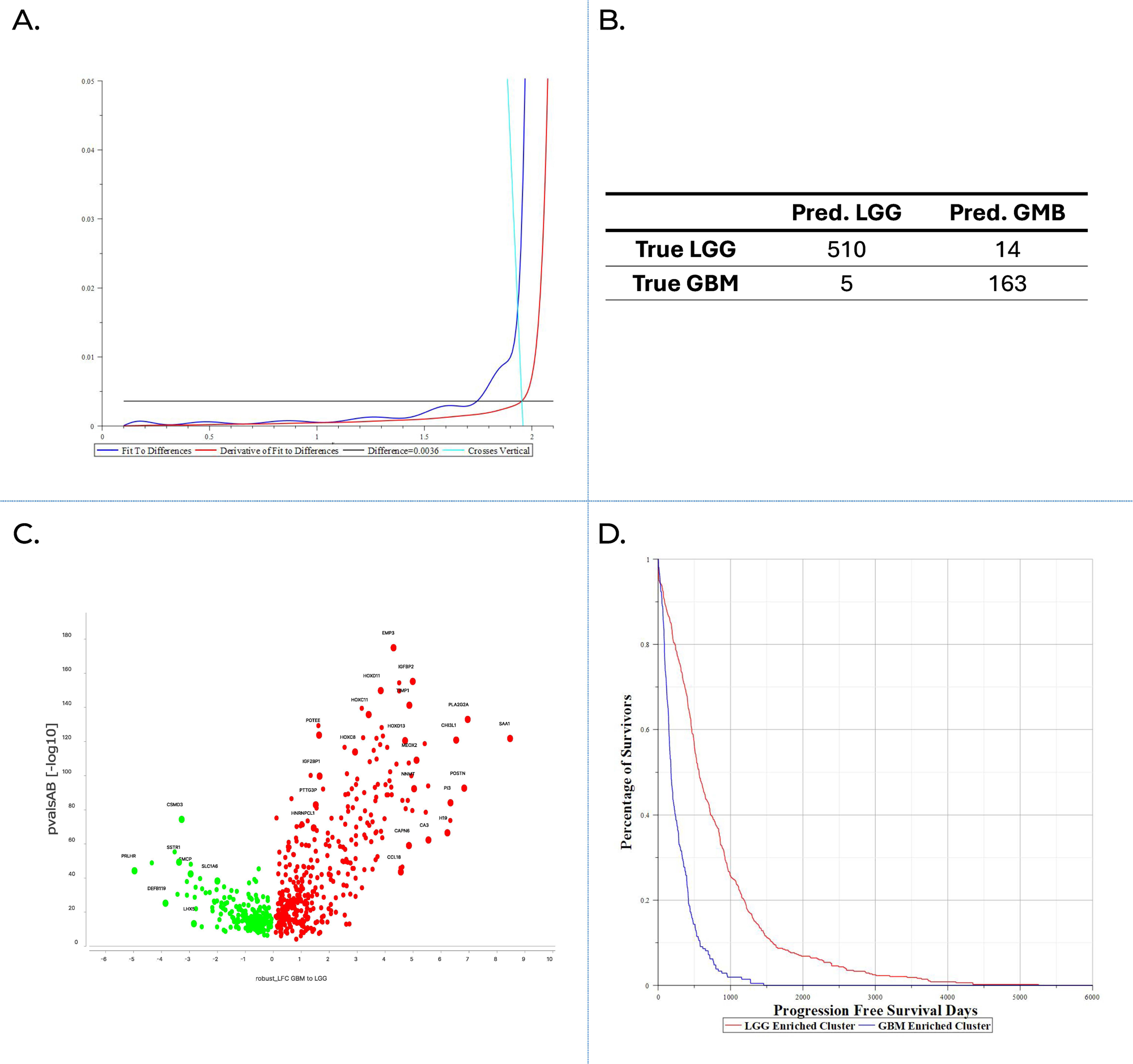
Denoising the transcriptome reveals a 554-gene core progression signal with stronger clinical separation. To identify the genes most responsible for separating the two major tumor states, all 20,057 protein-coding genes were ranked by the absolute difference in mean expression between the two full-transcriptome DQC groups. The elbow of this ranked curve defined the 554-gene cutoff, retaining the strongest separation signal while reducing weaker background variation. Histology and survival were not used for gene selection or clustering. (A) Selection of the 554-gene panel, with the fitted expression-difference curve shown in blue and its first derivative in red; the elbow marks the 554-gene cutoff. (B) Differential expression between the two 554-gene DQC states, showing genes enriched in the GBM-associated invasive and extracellular-matrix-remodeling state in red and genes enriched in the LGG-associated neurodevelopmental state in green; representative genes are labeled. (C) Post hoc comparison with TCGA histology, showing 97.3% concordance between the RNA-defined states and LGG or GBM diagnosis (673 of 692 tumors). (D) Kaplan–Meier analysis testing whether the RNA-defined states also differed in progression-free survival. The GBM-enriched invasive state had markedly shorter progression-free survival than the LGG-enriched neurodevelopmental state. Together, these panels show that reducing weaker transcriptomic variation preserved a core glioma progression signal while strengthening its association with diagnosis and patient outcome.

The 554-gene threshold corresponded to the elbow of the expression-difference curve, identified by a numerical first-derivative scan. In practical terms, this threshold marked the point where the strongest separation signal was retained while weaker, less informative variation from the protein-coding transcriptome was reduced. When DQC was applied to this 554-gene panel, the continuous glioma trajectory was preserved, but the tumors condensed into two additional stable endpoint groups (**Figure S4**).

We next tested whether this improved separation reflected a reproducible biological signal rather than a visualization artifact. Varying the underlying visualization dimensions produced near-identical tumor partitions, supporting the robustness of the 554-gene structure.

Having defined these two refined tumor states without using diagnostic labels, we then examined the biological programs that distinguished them. Differential expression analysis using Welch’s t-test with FDR < 0.05 identified strong transcriptomic differences between the two diagnostic-agnostic clusters (**Figure 3B**, **Table S2**). Cluster 2 showed marked up-regulation of extracellular matrix remodeling, invasion-related programs, including matrix metalloproteases, and pro-inflammatory signaling pathways, including IL-17 and NF-κB compared to Cluster 1. This invasive state was also characterized by a developmental HOX signal, including *HOXA5*, *HOXA6*, and *HOXD11*, as well as *EMP3* expression. In contrast, Cluster 1 retained more neurodevelopmental features, including higher expression of neuronal markers such as *NEUROD6* and *SPHKAP* (**Table S4**, **Figure S5**). IPA-curated pathway annotations are provided in **Table S3**.

Finally, we overlaid TCGA clinical metadata to determine whether these biologically distinct RNA-defined states had clinical relevance. Compared with the full-transcriptome baseline, the 554-gene model produced a more than three-fold reduction in mixed-label samples at the diagnostic boundary. The neurodevelopmental state, Cluster 1, was overwhelmingly LGG-enriched, containing 510 of 524 LGGs. The invasive, ECM-remodeling state, Cluster 2, was primarily GBM-enriched, containing 163 of 168 GBMs.

This label-free assignment achieved an overall diagnostic accuracy of 97.3%, corresponding to 673 of 692 tumors, with a positive predictive value of 99.0% for LGG and 92.1% for GBM (**Figure 3C**). Kaplan–Meier analysis showed that the GBM-enriched invasive state, Cluster 2, was associated with markedly shorter OS (p<1E-30) (**Figure 3D**).

Together, these results indicate that reducing background transcriptional noise strengthened the clinically relevant glioma signal. The 554-gene panel preserved the major LGG-to-GBM organization, improved histology concordance, and aligned strongly with patient outcomes, all without using diagnosis or survival information to build the clusters.

### 4. DQC shows that a compact 90-gene set preserves LGG–GBM separation and reveals distinct LGG risk groups

After defining the 554-gene RNA panel, we asked whether this signal could be reduced further into a more compact and potentially practical gene set. To identify the genes contributing most strongly to separation between the two primary tumor states, we ranked the 554 genes using a standard two-sample t-test. This test was used only as an objective ranking method to prioritize genes; it was not used to define statistical significance or test a biological hypothesis.

We then repeated DQC using progressively smaller sets of the highest-ranked genes, including the top 140, 100, and 90 genes (**Table S5**). Reducing the number of genes did not weaken the main tumor pattern. Instead, it revealed additional clinically meaningful groups.

At the 140-gene threshold, DQC separated the tumors into three groups: Cluster A with 450 tumors, Cluster B with 177 tumors, and Cluster C with 65 tumors. When clinical metadata were added afterward, Clusters A and C were composed entirely of LGGs, whereas Cluster B contained all 168 GBMs and 9 LGGs. Thus, 94.9% of the tumors in Cluster B were GBMs. Kaplan–Meier analysis of PFS showed significant differences among the three groups (two-sided log-rank *P* < 0.001). Cluster B represented the GBM-rich state with the poorest survival, with mean PFS/OS of 275/445 days. The two LGG groups showed distinct risk profiles, with mean PFS/OS of 811/1053 days for Cluster A and 390/660 days for Cluster C (**Tables S5-1-1** and **S5-1-2**).

Further reduction to the 100-gene threshold separated the tumors into four distinct groups: Cluster A with 421 tumors, Cluster B with 173 tumors, Cluster C with 65 tumors, and Cluster D with 33 tumors. When clinical information was added afterward, Cluster B contained all 168 GBMs and only five LGGs. Thus, 97.1% of the tumors in Cluster B were GBMs. Clusters A, C, and D contained only LGGs.

Kaplan–Meier analysis of PFS showed significant differences among the four groups (two-sided log-rank *P* < 0.001). Mean PFS/OS values were 835/1062 days for Cluster A, 448/1044 days for Cluster D, 397/602 days for Cluster C, and 263/430 days for Cluster B, preserving the survival order A → D → C → B (**Tables S5-2-1** and **S5-2-2**). This four-group pattern remained stable when the gene set was reduced to 90 genes. Clusters A, C, and D contained only LGGs, whereas Cluster B contained all 168 GBMs and only five LGGs. Thus, 97.1% of the tumors in Cluster B were GBMs. The same survival order was also preserved. Mean PFS/OS values were 836/1063 days for Cluster A, 458/871 days for Cluster D, 406/692 days for Cluster C, and 263/430 days for Cluster B, giving the survival order A → D → C → B (**Figure 4A–B**; **Tables S5-3-1** and **S5-3-2**). When fewer than 90 genes were used, this stable four-group pattern was no longer maintained [add the supporting supplementary figure or table citation here]. The 90-gene set was therefore the smallest tested gene set that preserved the main histologic and survival-related pattern.

**Figure 4.**
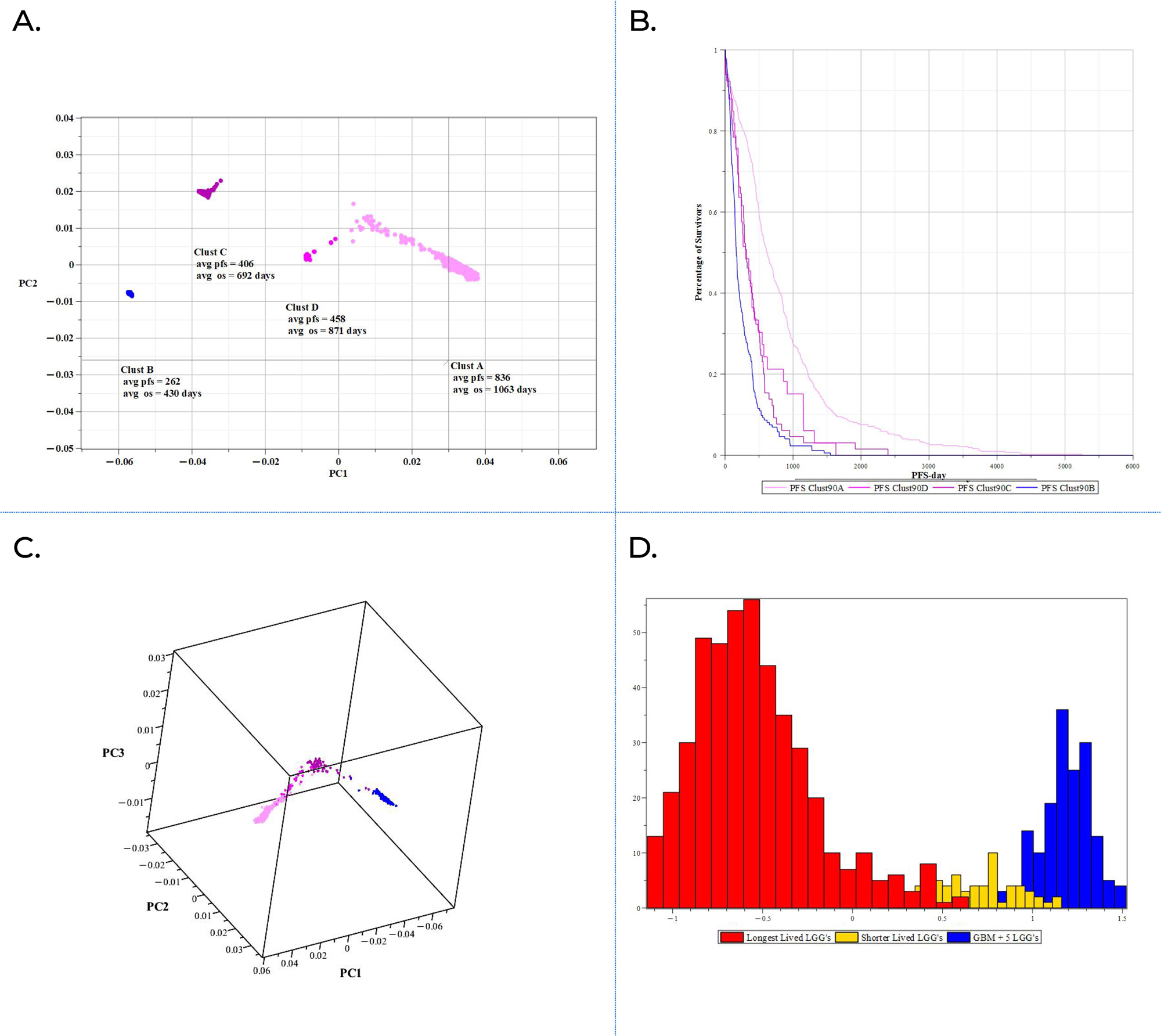
DQC separates gliomas into four risk groups using 90 genes, and broader RNA differences between these groups are used to create BioCoords. DQC was repeated with progressively smaller top-ranked gene sets, the 90-gene set was the smallest that preserved four stable tumor groups. Histology and survival were added only afterward. Selected group pairs were then compared across all 20,057 protein-coding genes, and the strongest RNA differences were combined into four biological scores, called BioCoords, for each tumor. (A) Final DQC map based on the 90-gene set, showing four RNA-defined tumor groups. After clinical information was added, Clusters A, D, and C contained only LGGs and showed progressively shorter mean survival, whereas Cluster B contained all 168 GBMs and five LGGs and had the poorest outcomes. (B) Kaplan–Meier analysis of progression-free survival across the four groups, confirming the survival order A → D → C → B (two-sided log-rank *P* < 0.001). (C) Final DQC view based on the four BioCoord scores, shown in three dimensions using the first three principal components. Each point represents one tumor. The preserved four-group pattern shows that the BioCoords retained the main tumor structure identified by the 90-gene analysis. (D)Distribution of tumors along the A–B mesenchymal–invasion BioCoord. Red indicates the longest-surviving LGGs, yellow indicates shorter-surviving LGGs, and blue indicates all GBMs together with the five LGGs assigned to Cluster B. Lower scores were seen mainly in the longest-surviving LGGs, whereas higher scores showed greater activity of the mesenchymal–invasion program and were concentrated in the GBM-enriched group. Together, these panels show how DQC identified four clinically distinct tumor groups from a compact gene set and how the broader RNA differences between those groups were converted into biological scores for individual tumors.

Together, these stepwise results show that the main glioma RNA pattern can be retained using a focused 90-gene set. This gene set preserves the LGG–GBM separation while also revealing stable LGG risk groups with distinct clinical outcomes, without using diagnosis or survival information to form the groups.

### 5. From four DQC-defined glioma groups to biological scores linked to patient outcome

DQC applied to the 90-gene set organized the tumors into four groups with different clinical outcomes: three LGG-only groups with progressively shorter survival in the order A, D, and C, and one GBM-enriched group with the poorest outcomes, Cluster B (**Figure 4A**; **Figure S6-1**). Survival information was added only after the groups had formed. Kaplan–Meier analysis confirmed this ordering, with mean PFS/OS values of 836/1063 days for Cluster A, 458/871 days for Cluster D, 406/692 days for Cluster C, and 263/430 days for Cluster B (**Figure 4B**).

We next asked whether the broader RNA differences among these four fixed groups could be converted into biological scores for individual tumors. We summarized these differences into four scores, called BioCoords. Unlike the four fixed groups, each BioCoord places every tumor along a range from lower to higher activity of a particular biological program.

### Creating BioCoords from RNA differences among the four glioma groups

The survival order A → D → C → B was used to guide which groups were compared. We compared A with B and D with B to capture differences between the LGG groups and the GBM-enriched group. We also compared A with C and C with D to capture differences among the LGG-only groups. The A–D and B–C comparisons were not retained because they did not add a clearly distinct RNA pattern beyond those already captured by the four selected comparisons.

Importantly, the genes used to create the BioCoords were selected from the full set of 20,057 protein-coding genes, not only from the 90-gene set. For each comparison, a two-sample *t*-test was used only to rank the genes by how strongly they differed between the two groups. The top 30 genes from each comparison produced 120 candidate genes. After repeated genes were assigned to only one gene group, 95 unique genes remained across four groups containing 24, 25, 27, and 19 genes (**Table S7**).

Because genes differ in their usual expression ranges, each gene was first scaled relative to its mean and variation across the TCGA cohort. The scaled values were then averaged within each gene group to calculate four BioCoord scores for every tumor. A score near 0 indicates average activity of that biological program across the cohort. Negative values indicate lower-than-average activity, whereas positive values indicate higher-than-average activity. DQC was then applied again using the four BioCoord scores. The major tumor organization identified by the 90-gene analysis reappeared, extending from the longer-surviving LGG region toward the GBM-enriched, poorer-outcome region (**Figure 4C**). This showed that the four BioCoords retained much of the RNA-defined tumor structure while making the biological differences underlying that structure easier to interpret.

### Four BioCoords link glioma organization and survival to invasion, development, metabolism, and cell maintenance

The genes defining the four BioCoords were not random collections. Each module contained genes associated with recognizable biological functions, giving biological vocabulary to the tumor organization revealed by DQC. Rather than describing each tumor through thousands of individual expression values, the BioCoords describe its relative position along four coordinated biological programs.

Among the four BioCoords, the A–B mesenchymal–invasion score showed the clearest relationship with clinical risk. Tumors with lower A–B scores showed lower relative expression of this program and were enriched for the longest-surviving LGGs. Tumors with intermediate scores included LGGs with shorter survival, whereas tumors with higher scores showed greater expression of the mesenchymal–invasion program and were concentrated mainly in the GBM-enriched, poorer-outcome region of the map (**Figure 4D**). The A–B BioCoord therefore converts the difference between the longest-surviving LGG group and the GBM-enriched group into a continuous biological score associated with increasing clinical risk.

The A–B BioCoord represents mesenchymal and invasion-related activity. This 24- gene module contains *ANXA2P2, CASP6, CLIC1, EMP3, GNG5, HOXA6, HOXB9, HOXC11, HOXC13, HOXC9, HOXD11, IGF2BP3, IGFBP2, MAGED4B, PABPC3, PAX3, PDPN, PLA2G2A, RAB42, SLC43A3, TGIF1, TIMP1, TUBA1C,* and *VIM*. The module includes genes associated with extracellular-matrix remodeling and invasion, including *EMP3, IGFBP2, PDPN, TIMP1,* and *VIM*, together with several HOX-related transcriptional regulators. Its graded relationship with histology and survival is shown in **Figure 4D**, where increasing scores were associated with movement from the longer-surviving LGG region toward the GBM-enriched, poorer-outcome region.

The A–C BioCoord represents development and cell identity. This 25-gene module contains *DCTD, EN1, EVX2, GNG12, HOXA-AS2, HOXA-AS3, HOXA1, HOXA2, HOXA3, HOXA4, HOXA5, HOXA7, HOXA10, HOXC4, HOXC6, HOXC10, HOXD9, HOXD10, HOXD13, ISL2, OSR2, OTP, PTGFRN, PYGL,* and *RAET1K*. The module is dominated by HOX genes and also includes neural-development and patterning genes such as *EN1, EVX2, ISL2,* and *OTP*. It therefore captures differences in developmental regulation and cell identity among the longer- and shorter-surviving glioma groups. Because the data represent single tumor samples rather than longitudinal measurements, this coordinate is best interpreted as distinguishing developmental states rather than proving that individual tumors progressively lose a particular lineage identity.

The D–B BioCoord represents mitochondrial, protein-maintenance, and neuronal programs. This 27-gene module contains *ARMC10, CHCHD2, CKS1B, COMMD10, EIF3CL, FAM226B, GUCA2A, HSP90B2P, KCNB1, LIMS3, MRPL36, MYH14, PGAM4, PIPSL, PPIA, PRELID1, RAP1B, RHAG, RPL13AP6, RPS3A, RPS7, SGCD, SLC25A21-AS1, SPAG4, TMEM183B, UQCRHL*, and *ZFAND2A*. These genes include components associated with mitochondrial function, protein translation and maintenance, and specialized neuronal or cellular features. This BioCoord indicates that the LGG and GBM-enriched regions differ not only in mesenchymal and invasion-related activity but also in fundamental cellular functions and the preservation of specialized cell identity.

The C–D BioCoord represents developmental and cell-maintenance programs. This 19-gene module contains *C1QTNF2, CCDC150, CCDC34, CDCA7L, CHAF1B, DNAJB12, DNAJC12, DNAJC2, HAP1, HOXA10-AS, HOXB3, HOXB7, HOXD12, LIG1, MDGA2, SKP2, SPOCK1, SUSD5*, and *ZNF473*. The module includes HOX-related developmental regulators, DNAJ-family chaperone and co-chaperone genes, genes involved in DNA replication and chromatin maintenance, and genes related to cellular adhesion. It mainly distinguishes the intermediate LGG groups, showing that LGGs with different outcomes also differ in coordinated developmental and cell-maintenance programs.

Together, these four BioCoords give biological meaning to the DQC map. The separation among the glioma groups was associated with coordinated RNA differences involving invasion and extracellular-matrix remodeling, developmental identity, mitochondrial and translational functions, protein maintenance, and cellular organization. By summarizing these programs as continuous scores, the BioCoords allow each tumor’s position in the DQC map to be interpreted biologically while preserving the broader organization from longer-surviving LGG states toward the GBM-enriched, poorer-outcome state.

### DQC reveals how progression-free survival aligns with mesenchymal and developmental glioma states

To examine how patient outcome related to this biological organization, we generated a survival animation from the original four-dimensional DQC analysis in BioCoord space and selected three representative frames for display (**Figure 5**). In each frame, every tumor is positioned horizontally according to its A–B mesenchymal– invasion and A–C developmental BioCoord scores, while its observed progression-free survival (PFS) is shown on the vertical axis (**Figure 5**).

**Figure 5.**
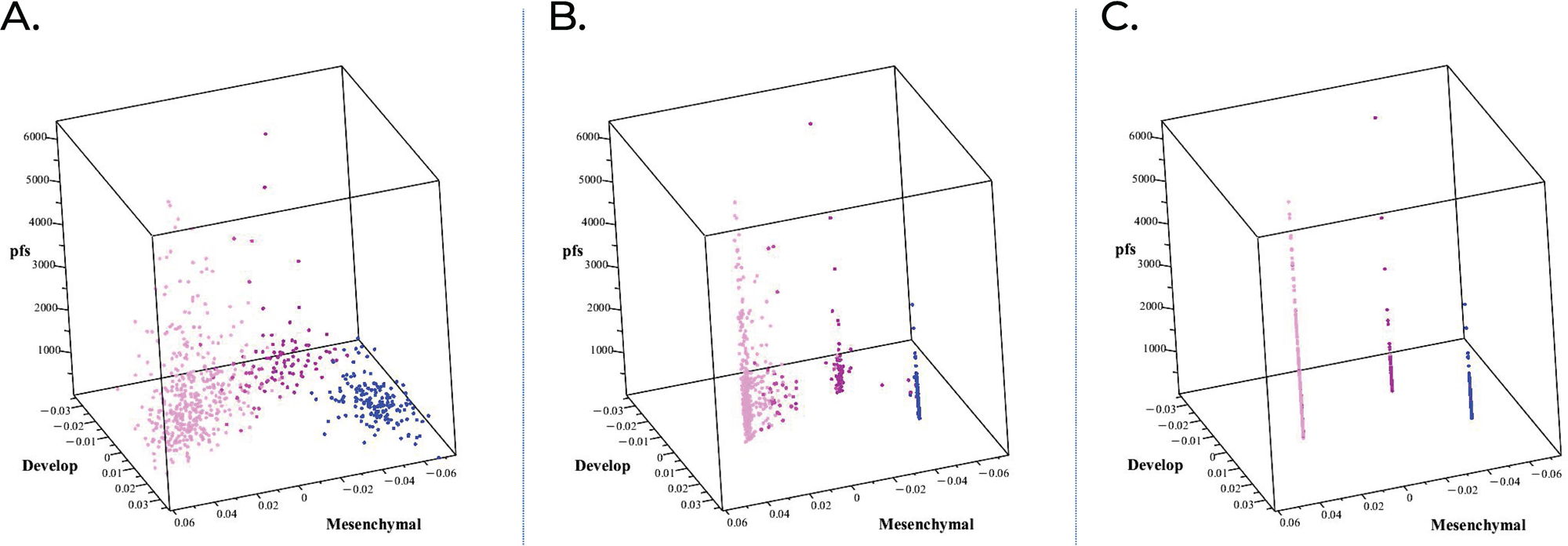
DQC animation reveals how progression-free survival aligns with mesenchymal and developmental glioma states. The panels show three frames from a survival animation built from the original four-dimensional DQC analysis of TCGA gliomas in BioCoord space. For each tumor, the mesenchymal–invasion and developmental BioCoord values are the same as in the original DQC animation, while the third, vertical coordinate shows that tumor’s observed progression-free survival (PFS). DQC therefore organizes tumors by biological similarity, while each tumor retains its own PFS as its vertical position. PFS did not guide DQC or influence group formation; it was added only to reveal how outcome varied with biological position. As DQC progressed, the tumors changed from a broad cloud to sheet-like survival patterns and finally to vertical survival structures above the DQC groups. Each point represents one tumor. TCGA histology was added afterward for interpretation: magenta, lower-grade glioma (LGG); blue, glioblastoma (GBM). (A) Starting DQC frame, showing tumors broadly distributed across both BioCoords and a wide range of PFS values. (B) Intermediate frame, showing biologically similar tumors drawing together while retaining their individual PFS values. The emerging sheets reveal that different combinations of mesenchymal and developmental activity are associated with different ranges of survival. (C) Final DQC frame, showing three prominent vertical survival structures visible in this two-coordinate view: two LGG-dominant structures with different biological positions and PFS distributions, and one compact GBM-enriched structure concentrated at shorter PFS. These vertical structures represent the distribution of PFS above the corresponding biological groups; they are not additional clusters. Together, these panels show that survival is systematically related to RNA-defined tumor biology and help explain why patients within the same broad diagnostic category can experience different disease courses.

PFS did not guide DQC and did not influence how tumors moved or how the groups formed. Instead, each tumor retained its recorded PFS value while DQC organized the tumors according to similarity across the four BioCoords. The visualization therefore displays three dimensions from the four-dimensional BioCoord analysis: two biological coordinates and PFS (**Figure 5**). This design allowed the relationship between RNA-defined biological position and clinical outcome to become visible during the DQC animation.

In the starting frame, the tumors formed a broad cloud with wide variation in both biological position and PFS, and the relationship between the biological map and survival was not yet clear (**Figure 5A**). As DQC progressed, tumors with more similar BioCoord profiles drew together while retaining their individual PFS values. Broad, sheet-like survival patterns then emerged above the biological map, showing that different combinations of mesenchymal and developmental activity were associated with different ranges of PFS (**Figure 5B**).

The relationship with survival was especially apparent along the mesenchymal– invasion direction. As tumors occupied regions with higher mesenchymal–invasion scores, the distribution of observed outcomes shifted toward shorter PFS. This pattern was systematic rather than absolute: nearby tumors often showed similar PFS distributions, but tumors occupying the same biological region did not necessarily have identical outcomes.

At the final frame, the survival sheets contracted into vertical structures above the DQC groups (**Figure 5C**). These vertical structures are not additional clusters. Each structure represents the distribution of individual PFS values among tumors occupying the corresponding biological region. Three prominent structures were visible in this two-coordinate view: two LGG-dominant structures with different mesenchymal and developmental positions and PFS distributions, and one more compact GBM-enriched structure concentrated at shorter PFS.

The persistence of a range of PFS values within each vertical structure is also biologically important. A DQC group was not associated with one fixed survival time. Some patients within a generally favorable biological region had shorter PFS, while variation remained even within the poorer-outcome region. Tumor RNA expression does not capture every factor affecting clinical outcome. Treatment history, immune activity, vascularization, the local tumor environment, coexisting medical conditions, and other unmeasured factors may contribute to differences among tumors with similar BioCoord profiles.

The vertical survival structures complement the Kaplan–Meier curves. Kaplan–Meier analysis summarizes survival across a group, whereas the vertical structures preserve the observed range and distribution of individual PFS values associated with each biological region. The LGG-dominant regions occupied a broader area of BioCoord space and contained wider PFS distributions. In contrast, the GBM-enriched region was more compact and concentrated mainly at shorter PFS.

Together, these findings show that PFS is systematically related to the combined activity of several RNA-defined biological programs rather than to histologic diagnosis or one biological score alone. At the same time, the remaining variation within each biological region shows that BioCoord position does not completely determine outcome. The BioCoord framework therefore provides an interpretable way to visualize why tumors carrying the same broad diagnosis can occupy different biological states and be associated with markedly different disease courses.

### 6. Cross-cohort validation in CGGA shows that BioCoords preserve clinically relevant glioma structure

To determine whether the BioCoord framework generalizes to independent clinical datasets, we analyzed 693 tumors from the Chinese Glioma Genome Atlas (CGGA). This cohort comprised 443 lower-grade gliomas (LGG; 282 primary, 161 recurrent) and 249 glioblastomas (GBM; 140 primary, 109 recurrent) (**Fig S7A**). For the cross-cohort DQC validation, tumors were then restricted at the outset to the 657 CGGA cases with available overall-survival and clinical annotation data. The CGGA dataset differs substantially from TCGA in both gene coverage and preprocessing pipelines. To test the framework under these cross-cohort differences, we restricted the analysis to the 16,154 genes shared between the two platforms. As a result, only 78 of the 95 original BioCoord core genes were retained in the validation cohort. This provided a stringent test of whether the BioCoord framework captures reproducible glioma biology rather than a TCGA-specific expression pattern.

### Direct gene-expression clustering did not robustly recover prognostic structure

Applying the label-free DQC algorithm directly to the 16,154 shared genes yielded a diffuse, unstructured organization. Although the algorithm induced local sample movement, LGG and GBM tumors remained substantially overlapping (**Figure S8**).

Post hoc survival analysis confirmed these unstructured gene-space groupings did not correspond to meaningful differences in overall survival (**Figure 6A**).

**Figure 6.**
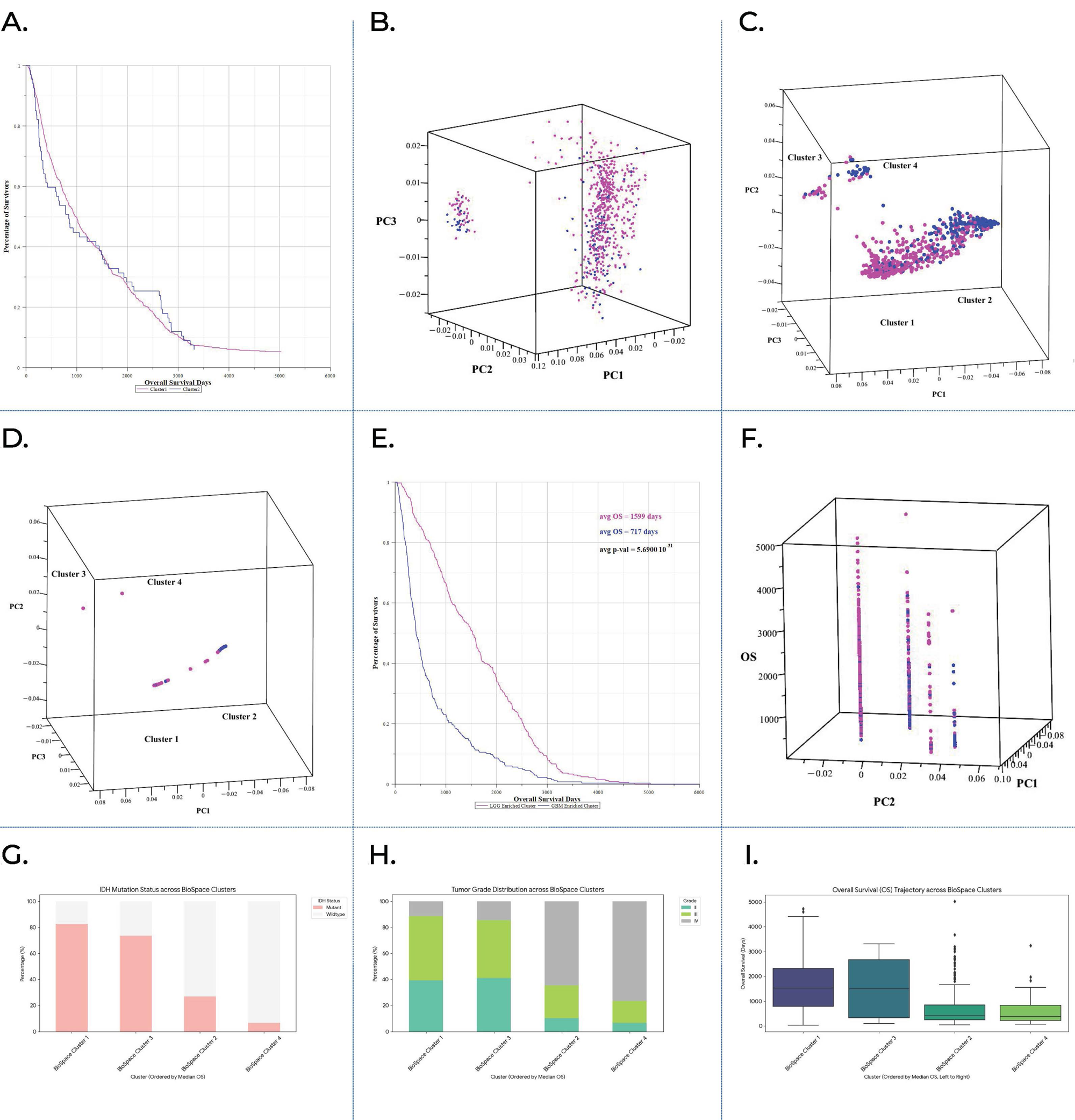
BioCoord representation recovers clinically informative glioma structure in the independent CGGA cohort. BioCoords were calculated from CGGA RNA expression for 657 tumors with available overall-survival data using retained TCGA-defined gene modules; clinical, molecular, treatment, and survival annotations were added only after clustering and are provided in Supplementary Table S7. (A) Direct shared-gene clustering. Kaplan–Meier analysis of the two major DQC clusters derived from the 16,154 shared TCGA–CGGA genes showed no clear overall-survival separation among CGGA tumors with available OS data, indicating that direct gene-expression clustering did not recover prognostic structure in the validation cohort. (B) Knee-based feature selection. Knee-based denoising reduced the CGGA gene-expression matrix to approximately 636 genes and produced visually clearer grouping, but LGG and GBM samples still remained substantially overlapping after post hoc diagnostic overlay. (C–D) BioCoord projection of CGGA tumors. CGGA tumors mapped with TCGA-derived BioCoords formed an organized LGG-to-GBM-like structure. Histology was added only afterward, yet LGG and GBM tumors occupied different regions of the map rather than mixing randomly. This suggests that BioCoords capture clinically relevant tumor-state differences that are preserved across cohorts. (E) Post hoc survival separation. Kaplan–Meier analysis showed longer overall survival in BioCoord-derived cluster 1 than cluster 2 among OS-available CGGA tumors. (F) Survival distribution within BioCoord space. Overall survival plotted directly in BioCoord space showed that longer-surviving cases were enriched in the LGG-dominant region, whereas shorter-surviving cases were enriched toward the GBM-enriched region, supporting a survival-associated spatial organization within the BioCoord map. (G) IDH status across BioCoord clusters. IDH status differed across the BioCoord-defined tumor groups. Clusters with longer survival were enriched for IDH-mutant tumors, whereas shorter-survival clusters were enriched for IDH-wildtype tumors. (H) WHO grade across BioCoord clusters. WHO grade also differed across the BioCoord-defined tumor groups. Longer-survival clusters contained higher proportions of Grade II/III tumors, whereas shorter-survival clusters were enriched for Grade IV tumors. (I) Overall survival across BioCoord clusters. OS distributions differed across BioCoord-defined tumor groups ordered by median OS, with clusters 1 and 3 showing longer survival and clusters 2 and 4 showing shorter survival. Together, these analyses show that BioCoord-based organization captures clinically meaningful glioma structure in CGGA that was not recovered by direct gene-expression clustering or feature selection alone.

We next applied multiple objective feature-selection strategies, including a knee-based variance filter retaining approximately 636 genes and p-value-based rankings, to test whether stronger gene filtering could improve clinical separation. These reduced gene sets produced clearer visual clustering and local contraction (**Figure 6B**, **Figure S9**), but they still showed limited prognostic separation. In addition, standard statistical rankings did not consistently retain several genes contributing to the TCGA BioCoord axes, including HOX-related developmental programs. These findings suggest that direct gene-level clustering is sensitive to cohort-specific differences in preprocessing, gene coverage, and feature selection, making it difficult to recover the same survival-associated tumor organization across datasets.

### BioCoord-derived tumor groups align with clinical outcome across cohorts

We next tested whether the TCGA-derived BioCoord framework could recover clinically meaningful tumor organization in the independent CGGA cohort. Of the 95 TCGA-defined BioCoord genes, 78 were retained in CGGA and mapped to their corresponding TCGA-defined modules. BioCoord values were calculated for each CGGA tumor using CGGA RNA expression, and DQC was then applied to these four BioCoord values. Diagnostic labels, molecular annotations, and survival data were added only after BioCoord calculation and clustering.

In contrast to direct gene-expression clustering, BioCoord-based analysis produced a more organized CGGA tumor map (**Figure 6C–D**, **Figure S10**). At the intermediate DQC frame, most tumors formed a main continuous structure, with smaller separated groups nearby. When histology was added afterward, LGG-labeled tumors were enriched toward one side of the main structure and GBM-labeled tumors toward the other, showing an LGG-to-GBM-like organization rather than random mixing (**Figure 6C**; **Figure S10**). At a later frame, the map contracted into a tighter trajectory-like pattern while preserving the relative positions of the separated tumor groups (**Figure 6D**; **Figure S10D–E**). By the final DQC frame, this BioCoord map resolved into four spatially separated endpoint clusters, providing discrete tumor groups for post hoc clinical evaluation (**Figure S10F**).

Post hoc survival analysis showed that these final BioCoord-derived clusters were associated with distinct overall-survival outcomes in CGGA. In the main comparison, Cluster 1 showed longer survival than Cluster 2, with average OS of 1599 versus 717 days (**Figure 6E**). Integration of the final DQC/BioCoord cluster assignments with CGGA clinical metadata further showed that cluster 1 contained 328 tumors and had a median OS of 1530 days, with enrichment for LGG, IDH-mutant, and 1p/19q- codeleted tumors. In contrast, cluster 2 contained 265 tumors and had a median OS of 414 days, with enrichment for GBM, IDH-wildtype, and non-codeleted tumors (**Table S7**). In the supplementary comparison, cluster 3 showed longer survival than cluster 4, with average OS of 1570 versus 659 days (**Figure S11D**). Importantly, when OS was plotted directly within BioCoord space, survival was not randomly distributed. Instead, tumors aligned along survival-associated trajectories within the BioCoord map, with longer-surviving cases enriched in LGG-dominant regions and shorter- surviving cases enriched toward GBM-enriched regions (**Figure 6F**). This pattern was consistent with the spatial organization observed across the BioCoord DQC frames, where tumors moved from broad, continuous structures toward ordered endpoint groups (**Figure 6C–D**; **Figure S10B–F**; **Figure S11C**). Together, these plots suggest that BioCoord position captures both tumor identity and outcome-related progression structure.

This survival-associated structure was not recovered by direct clustering of the 16,154 shared genes or by knee-based feature selection alone (**Figure 6A–B**). Thus, the BioCoord framework preserved outcome-associated tumor organization in an independent cohort despite differences in gene coverage, preprocessing, and sample composition.

To determine whether the BioCoord-defined groups corresponded to clinically recognizable tumor features, we next examined their molecular and histologic composition using the CGGA annotation variables summarized in **Table S7**. Longer-survival BioCoord groups were enriched for IDH-mutant tumors, whereas shorter-survival groups were enriched for IDH-wildtype tumors (**Figure 6G**; **Table S7**). WHO grade distribution also shifted across the BioCoord clusters: longer-survival groups contained a higher proportion of lower-grade gliomas, whereas shorter-survival groups were enriched for Grade IV tumors (**Figure 6H; Table S7**). Consistent with these patterns, overall-survival distributions differed across the ordered BioCoord groups, with clusters 1 and 3 showing longer survival distributions and clusters 2 and 4 showing shorter survival distributions (**Figure 6I; Table S7**).

Together, these findings show that BioCoord-derived tumor groups in CGGA align with multiple clinically relevant features, including histology, WHO grade, IDH status, treatment context, and overall survival. Importantly, the BioCoord groups did not simply reproduce one clinical annotation. Instead, they summarized coordinated RNA-expression programs that tracked with several features used in real-world glioma diagnosis and prognosis.

To place these findings in the context of established clinical and molecular predictors, we examined overall survival across standard CGGA annotations (**Figure S12; Table S7**). Survival decreased with increasing WHO grade, while IDH mutation and 1p/19q codeletion were associated with longer survival. MGMT promoter methylation showed a more modest survival difference. However, within WHO Grade III tumors, survival differed substantially by IDH status, showing that histologic grade alone did not fully capture outcome heterogeneity.

We then examined whether these survival patterns were consistent across age and treatment contexts (**Figure S13; Table S7**). Survival differences associated with WHO grade, IDH mutation, MGMT promoter methylation, and 1p/19q codeletion were generally preserved, but their magnitude varied across strata. Survival was shorter in older patients and further reduced among older untreated patients, even when tumors were grouped by the same diagnostic or molecular feature. These analyses show that established markers remain clinically informative, but their prognostic impact depends on clinical context.

Overall, the CGGA validation shows that BioCoords provide a more transferable RNA-based tumor map than direct gene-expression clustering alone. In this independent cohort, BioCoord-derived organization aligned with histologic diagnosis, WHO grade, IDH status, treatment context, and overall survival.

Importantly, survival was not only different between endpoint clusters; it also followed the spatial organization of tumors within BioCoord space, supporting the idea that BioCoords capture outcome-associated progression structure rather than simply reproducing standard clinical labels. These findings support BioCoords as an interpretable framework for connecting large-scale RNA data with real-world diagnostic and prognostic questions in glioma.

## Discussion

Large public RNA-seq datasets contain a vast amount of biological and clinical information, but much of this information remains difficult to use in practice. The central challenge is not only that the data are large, but that thousands of gene-expression measurements must be converted into interpretable disease maps that researchers, pathologists, oncologists, and translational teams can understand and use [56,57]. With appropriate validation, these maps could help health professionals navigate patient care, assess disease progression and treatment response, and guide biomarker discovery and therapeutic development. We used adult-type diffuse glioma as a test case for this broader problem because glioma diagnosis already depends on integrated histologic and molecular criteria, yet clinically important outcome heterogeneity persists within standard diagnostic categories[14,15,56]. The 2021 WHO classification reflects major progress in incorporating molecular features into adult diffuse glioma diagnosis, including IDH status and 1p/19q codeletion, but it also highlights the need for tools that capture additional biological variation relevant to prognosis and progression [5,58].

In this study, we show that RNA expression alone can reveal a clinically meaningful organization of glioma without using histology, progression-free/overall survival, or other clinical features during the discovery process. In TCGA, protein coding transcriptome DQC organized tumors into LGG-enriched and GBM-enriched states that aligned strongly with post hoc diagnostic labels and survival. This finding suggests that RNA expression contains active biological information about tumor state, rather than simply reflecting labels assigned by human classification systems. In practical terms, the analysis converted a large gene-expression matrix into a readable tumor map: one that organized tumors by biological similarity and only afterward was shown to align with diagnosis and outcome.

A major lesson from the TCGA analysis is that the clinically relevant signal became clearer after reducing background transcriptional variation. Starting from more than 20,000 protein-coding genes, the workflow identified a 554-gene core progression signal that preserved the LGG-to-GBM-like organization while improving diagnostic concordance and survival separation. Further refinement to approximately 90 genes preserved the main LGG/GBM boundary and revealed distinct LGG risk groups with different clinical outcomes. This is important for real-world glioma care because lower-grade gliomas are not clinically uniform. Some remain relatively indolent, whereas others progress earlier or behave more aggressively despite sharing the same broad diagnostic label [14,15,58]. Earlier TCGA work also showed that molecular features such as IDH status and 1p/19q codeletion better capture clinically meaningful glioma structure than histology alone, reinforcing the value of molecular disease-state mapping [5,15,59–64].

Higher-risk states showed increased activity of extracellular matrix remodeling, invasion-related programs, inflammatory signaling, and mesenchymal behavior, consistent with a shift toward a more mesenchymal and invasive tumor state [18,64–67]. These recent longitudinal findings are particularly relevant because they show that mesenchymal-like states can increase during progression or recurrence and may be shaped by interactions between malignant cells and the surrounding immune environment. They also showed HOX-associated developmental reprogramming, suggesting that changes in developmental and lineage-related programs form another important axis of glioma progression [69–71]. These studies show that glioma progression can involve expansion of less differentiated or progenitor-like tumor-cell states, loss of more differentiated lineage programs, and changes in the epigenetic control of cell identity. Prior TCGA-based glioblastoma studies identified transcriptional subtypes, including mesenchymal and proneural patterns, supporting the idea that RNA-defined tumor states carry biological and clinical meaning [67,72,73]. In our study, these programs were not imposed during clustering; they were recognized only after the RNA-derived structure was generated. This distinction is central to the work: the goal was not to force tumors into known categories, but to allow the RNA data to organize first and then ask what biological programs explained the resulting map.

To make this tumor map easier to interpret and compare across datasets, we translated the 90-gene tumor-state structure into four BioCoords. Each BioCoord summarizes a coordinated biological program rather than relying on a single gene or a set of biologically unrelated individual markers. The dominant A–B BioCoord captured a mesenchymal and invasion-associated program, consistent with movement toward a more aggressive mesenchymal tumor state [17,18,24,60,65,73,74]. Another major coordinate captured a developmental axis marked by changes in developmental identity and lineage-related programs [61.62.68,75]. The remaining coordinates reflected metabolic and translational disruption, protein-homeostasis changes, and cell-maintenance programs. Together, these axes suggest that glioma progression is not driven by one isolated molecular event. Instead, it appears to reflect coordinated changes across several biological systems. This interpretation is consistent with recent single-cell studies showing that glioblastomas contain multiple interacting layers of biological variation, including malignant-cell states, tumor-wide expression programs, and distinct immune and stromal environments [24,75].

This multi-program view helps explain why prognosis is difficult to capture using one label or one molecular marker alone. In BioCoord space, tumors did not behave as isolated categories. They occupied a progression-related pattern in which survival varied across combinations of biological programs. LGG-like tumors showed broader biological and outcome variability, whereas GBM-enriched states were concentrated in poorer-outcome regions of the BioCoord space. This pattern matches a common clinical challenge: tumors with the same histologic or molecular diagnosis can still differ in aggressiveness, recurrence risk, and survival [14,15,72,75]. Recent large-scale single-nucleus analyses further show that individual glioblastomas contain different combinations of malignant-cell states and surrounding cell populations rather than conforming to one fixed biological category [75]. BioCoords provide a way to describe these differences as tumor-state patterns rather than as disconnected labels.

The CGGA validation was a stringent test of whether this framework captured disease-relevant biology rather than a TCGA-specific pattern. CGGA differed from TCGA in gene coverage, preprocessing, and cohort composition, and only 78 of the 95 TCGA-defined BioCoord genes were available. Because the validation analysis required outcome assessment, the cross-cohort analysis was performed on the 657 CGGA tumors with available overall-survival data; however, survival information was still used only after RNA-based BioCoord calculation and clustering. Direct clustering of the 16,154 shared genes did not recover meaningful survival separation in CGGA. Knee-based feature selection made the CGGA plots visually cleaner, but this improvement did not translate into clear diagnostic or prognostic separation. This result highlights an important translational point: a visually organized plot is not enough. For oncology and diagnostic use, the key question is whether the resulting tumor groups help explain diagnosis, prognosis, or biology.

In contrast, applying the TCGA-derived BioCoords to CGGA recovered tumor organization that was clinically meaningful. Even though only 78 of the 95 TCGA-defined BioCoord genes were available in CGGA, the BioCoord-defined groups aligned with overall survival, LGG/GBM diagnosis, WHO grade, and IDH status. Groups with longer survival were enriched for lower-grade and IDH-mutant tumors, whereas groups with shorter survival were enriched for Grade IV and IDH-wildtype tumors. Importantly, when overall survival was added afterward, longer- and shorter- surviving tumors occupied different regions of the BioCoord map. This suggests that the position of a tumor in BioCoord space reflects clinically relevant risk, not only membership in a final cluster.

These CGGA results support a practical point: BioCoords appear to capture coordinated tumor-state patterns that are preserved across datasets better than direct gene-level clustering alone. This matters because clinical glioma care depends on integrating many features, and no single feature fully explains patient outcome [5,14,64]. In CGGA, survival decreased with increasing WHO grade, while IDH mutation and 1p/19q codeletion were associated with longer survival. MGMT promoter methylation showed a more modest survival difference in this cohort, consistent with its established role as a marker of benefit from temozolomide treatment in glioblastoma [64].

However, these standard variables did not fully explain outcome heterogeneity. Within WHO Grade III tumors, IDH status separated patients into groups with different survival, showing that histologic grade alone was not sufficient. Age and treatment context also changed the apparent survival effect of standard markers. Together, these findings reinforce that glioma prognosis reflects the combined influence of tumor biology, molecular status, patient context, and treatment history. This framework should therefore be viewed as a complement to current glioma classification, not a replacement for it. Histology, IDH status, 1p/19q codeletion, MGMT promoter methylation, imaging, age, extent of resection, and treatment history remain essential for clinical decision-making. The added value of BioCoords is that they provide an interpretable RNA-based summary of tumor state. In future studies, this type of map could help researchers and clinicians identify higher-risk tumors within broad diagnostic categories, compare primary and recurrent tumors, and test whether tumors shift toward more aggressive biological states over time.

Recent analyses of paired initial and recurrent tumors demonstrate that glioma evolution can involve changes in malignant-cell states, immune and stromal composition, genetic alterations, and epigenetic regulation, supporting the need for frameworks that can follow several interacting dimensions of progression [58,60,66,75]. These applications require further validation, but the current results show that clinically relevant risk information can be recovered from RNA-seq data in a form that is interpretable across cohorts.

Although this study focuses on glioma, the larger idea is not limited to cancer. Many human diseases are shaped by coordinated biological programs rather than a single gene, pathway, or clinical label. Autoimmune diseases, neurodegenerative disorders, chronic inflammatory syndromes, infection-related illnesses, metabolic disorders, and complex multisystem conditions may all involve changes in biological state over time [76–78]. RNA expression provides a snapshot of what cells and tissues are actively doing. A BioCoord-like framework could therefore be useful for organizing large molecular datasets into disease-state maps that reflect severity, progression, treatment response, or disease mechanisms. This broader application will require careful testing, but it reflects the central goal of this work: to use computational tools as translators between large molecular datasets and real biological or medical questions.

Several limitations should be acknowledged. First, this study used retrospective bulk RNA-seq datasets from TCGA and CGGA. Bulk RNA-seq reflects a mixture of malignant cells, immune cells, stromal cells, vascular elements, and other microenvironmental signals, and the abundance and state of these components may differ substantially across regions of the same tumor [24,75,79,80]. This is clinically relevant because diagnostic tissue specimens are also mixed biological samples, but it limits our ability to assign each BioCoord program to a specific cell type. Single-cell, single-nucleus, and spatial transcriptomic studies will therefore be needed to determine which malignant and microenvironmental compartments contribute to each coordinate and whether their contributions vary across different regions of a tumor [24,75,79]. Second, CGGA validation was constrained by gene overlap; only 78 of the 95 TCGA-defined BioCoord genes were retained. The fact that the BioCoord structure remained clinically informative despite this limitation is encouraging, but additional validation in uniformly processed cohorts is needed. Third, clinical labels and survival data were added only after the RNA-based tumor structure was defined. This protects the discovery process from label-driven bias, but it also means that the survival findings are associations, not proof of causality. Treatment differences, tumor purity, sampling location, recurrence status, and unmeasured genetic, epigenetic, or microenvironmental features may contribute to the observed outcome patterns [60,66,79,81]. Fourth, this study does not yet establish independent clinical utility beyond standard predictors. Future studies should test whether BioCoords add prognostic information beyond WHO grade, IDH status, 1p/19q codeletion, MGMT promoter methylation, age, treatment, and extent of resection using multivariable models and prospective clinical cohorts.

Future work should focus on both biological validation and clinical translation. The BioCoord modules should be tested in additional glioma datasets with harmonized RNA-seq, methylation, imaging, treatment, recurrence, and longitudinal follow-up data. The mesenchymal/invasion and developmental-lineage axes should be studied experimentally to determine whether they reflect targetable mechanisms of progression, recurrence, or therapy resistance. Recent longitudinal and multimodal studies provide strong starting points by linking progression to mesenchymal-like changes, altered interactions with myeloid cells, expansion of less differentiated malignant-cell states, and changes in the genetic and epigenetic control of cell identity [60,62,66,75]. It will also be important to test whether smaller RNA panels can reproduce the BioCoord scores in clinically realistic settings. If validated prospectively, this type of framework could help identify higher-risk tumors within broad diagnostic categories, compare primary and recurrent tumors, and monitor whether a tumor is shifting toward a more aggressive biological state.

In summary, this study shows that label-free analysis of glioma RNA expression can recover a clinically meaningful disease-state map, refine it into a compact gene signature, and translate it into interpretable BioCoords. Across TCGA and CGGA, BioCoords preserved tumor organization linked to diagnosis, molecular status, and survival more effectively than direct gene-expression clustering alone. More broadly, this work supports a practical vision for big-data biology: computational methods should help researchers and clinicians convert complex molecular measurements into interpretable maps of disease state, pathogenesis, and patient risk. In glioma, and potentially in many other complex human diseases, such maps may help bring large-scale molecular data closer to the bedside.

## Supporting information

DQC Evolution Dynamics

## Data Availability

All data is available at:
https://www.cancer.gov/tcga
https://www.cgga.org.cn/
Software used in the analyses is available at:
https://github.com/zanderteller/dqm

https://github.com/zanderteller/dqm

https://www.cgga.org.cn/

## Resource availability

Software availability: https://github.com/zanderteller/dqm

## Acknowledgments

The results here are in whole or part based upon data generated by the TCGA Research Network: https://www.cancer.gov/tcga.

Research for the study was conducted under the 4C program at Stanford University. Funding provided by start-up funds from the University of Wisconsin-Madison.

## Declaration of generative AI and AI-assisted technologies in the writing process

During the preparation of this work, AI technologies were used to identify areas of the manuscript which were unclear and the authors subsequently rewrote sections of the manuscript to improve clarity.

## Supplemental Information

### Supplemental Notes

#### Supplemental Note S1-1. Intuitive description of Dynamic Quantum Clustering (DQC)

The TCGA glioma analysis revealed that extended structures emerging during DQC evolution, even when applied to noisy high-dimensional expression data, correspond to clinically meaningful patterns such as progression-free survival (PFS) and overall survival (OS). To help readers understand why this occurs, we provide an intuitive explanation of the DQC procedure using a simple two-dimensional example. A full mathematical description of the algorithm is available in the original publications ^1,2^.

The explanation below avoids mathematical formalism and instead illustrates the geometric intuition underlying the algorithm.

#### Supplemental Note S1-2. Model dataset used for illustration

To illustrate how DQC reveals hidden structure in complex data, we generated a two- dimensional dataset intended to mimic simplified RNA-seq measurements for two genes. The dataset contains four components:

1. **Three compact Gaussian clusters**, each representing a group of tumors with similar expression values. Differences among points within each cluster arise only from statistical variability.
2. **One extended distribution**, constructed by concatenating many overlapping Gaussian distributions arranged along a vertical trajectory. This extended structure represents a set of tumors whose gene expression changes smoothly along an underlying biological axis (for example, disease progression).

The combined dataset therefore contains both compact clusters and continuous structure, similar to patterns commonly observed in transcriptomic data.

**Figure S1** shows the simulated dataset.

#### Supplemental Note S1-3. Construction of the DQC potential function

DQC begins by constructing a potential function from the data.

This function can be understood as a smooth energy-like landscape defined across the data space.

Importantly:

● Regions containing many nearby samples correspond to lower values of the potential function.
● Regions with few samples correspond to higher values.

In intuitive terms, the potential surface highlights areas where samples accumulate.

To visualize this surface, the potential is plotted as a three-dimensional surface above the two-dimensional data plane. The third dimension does not represent an additional data variable; it simply displays the value of the potential function. For visualization, the hyperbolic tangent of the potential (tanh(V)) is shown to suppress large amplitudes away from the data and emphasize the topology near dense regions.

**Figure S2** illustrates this potential surface together with the original data points. In this representation:

● Compact clusters appear as circular depressions in the landscape.
● Sparse regions may contain multiple small local minima reflecting sampling noise.
● Extended data structures generate elongated valleys in the potential surface.

#### Supplemental Note S1-4. Evolution of samples during DQC

After constructing the potential surface, DQC evolves the positions of the data points according to a rule derived from quantum mechanical dynamics (the Hamiltonian). In practice, this rule simply determines how each point moves across the potential landscape during the algorithm.

An intuitive way to understand this process is to imagine each data point moving across the landscape while being influenced not only by its immediate surroundings but also by the broader structure of the landscape.

Two properties distinguish this evolution from classical gradient descent:

### 1. Non-local influence

Each point responds to a weighted average of the potential in its neighborhood rather than only the local slope at a single location. As a result, motion reflects the broader geometry of the data rather than small fluctuations in the surface.

### 2. Reduced sensitivity to small local minima

Because the evolution averages information across nearby regions, very small local depressions in the potential become effectively invisible. This prevents the algorithm from fragmenting data into many artificial micro-clusters, a common issue in high-dimensional datasets.

Together, these properties allow DQC to reveal large-scale structures such as extended trajectories or major clusters that reflect the intrinsic organization of the data.

### Supplemental Note S1-5. Example of DQC evolution

Figure S3 shows successive frames from a DQC animation applied to the model dataset. The red points represent the data samples evolving along the potential surface.

Key observations include:

● Initially separated clusters move toward stable positions within their respective valleys.
● The extended distribution gradually contracts along its principal axis.
● Small local irregularities in the potential do not trap points because the evolution responds to the broader landscape.

By later frames, the extended distribution collapses into a narrow linear structure representing a single dominant underlying axis in the data. This example illustrates how DQC can reveal hidden low-dimensional structure embedded within complex datasets. The full animation can be seen in this file (**DQC_Evolution_Dynamics**).

## Supplementary Figures

**Figure S1. Simulated dataset used to illustrate DQC behavior.**

Two-dimensional model dataset composed of three compact Gaussian clusters and one extended distribution generated from overlapping Gaussian components arranged along a vertical trajectory. The simulated data are used to illustrate how DQC distinguishes compact groupings from continuous structure.

**Figure S2. Potential function generated from the model dataset.** The DQC potential function is visualized as a three-dimensional surface above the two-dimensional data plane. Lower regions correspond to areas containing higher densities of samples. Data points are shown in red.

**Figure S3. Evolution of the model dataset during DQC.**

Panels S3-1 to S3-8) Selected frames from the DQC animation showing how samples move across the potential surface over the course of the algorithm. Compact clusters converge to stable basins, whereas the extended distribution contracts into a lower-dimensional structure that reflects the dominant organization of the data.

**Figure S4. DQC evolution of the 554-gene panel from diffuse cloud to two final clusters.**

Representative frames from Dynamic Quantum Clustering (DQC) applied to the 692×554 expression matrix, displayed in the first three principal components for visualization only. **Left**, frame 1: the initial diffuse cloud shows separation of the two dominant sample groupings. **Middle**, frame 11: the cloud contracts onto a lower-dimensional manifold while preserving the global LGG–GBM organization. **Right**, final frame: the data resolve into two compact endpoint clusters corresponding to the LGG-enriched and GBM-enriched states identified in the main analysis. Histology labels were overlaid only after clustering for post hoc evaluation.

**Figure S5. Pathway enrichment highlights ECM remodeling, inflammation, and invasion programs distinguishing DQC-defined clusters.** Ingenuity Pathway Analysis (IPA) of differentially expressed genes between the DQC-defined GBM-enriched and LGG-enriched clusters (FDR < 0.05) identified significant enrichment of extracellular matrix (ECM) organization and degradation pathways, including matrix metalloprotease activity and collagen-related processes. Additional enriched pathways include leukocyte adhesion and diapedesis, vascular transmigration, PI3K-associated motility, and pro-inflammatory signaling (e.g., IL-17, NF-κB, and PI3-family pathways). Pathways are ranked by Benjamini–Hochberg– adjusted p values. These results provide biological context for the unsupervised DQC separation, indicating that the GBM-enriched cluster is associated with ECM remodeling, invasion, and inflammatory signaling, whereas the LGG-enriched cluster retains more neuronal and developmental features.

**Figure S6. Compression of the 90-gene DQC structure into biological coordinates for risk stratification DQC-based clustering using the 90-gene signature.** An unsupervised reduction of the 554-gene set, driven by rank-ordering of genes by between-cluster contrast, yielded a 90-gene panel. Dynamic Quantum Clustering (DQC) resolved tumors into four clusters (A–D) without the use of clinical labels. While the GBM-enriched Cluster B notably shows five LGG samples that cluster biologically with the GBM group, suggesting an aggressive molecular phenotype. The clusters occupy an elongated trajectory (A→D→C→B) consistent with a dominant mesenchymal/invasion-associated gradient.

**Supplementary Figure S7. Clinical and histologic heterogeneity of the CGGA validation cohort.** CGGA clinical metadata were used to summarize the histologic composition of the independent validation cohort. The cohort included 693 diffuse gliomas spanning astrocytic lineage, oligodendroglial lineage, mixed/legacy oligoastrocytic lineage, and glioblastoma lineage, with one tumor lacking histology/grade annotation. Within each lineage, tumors were subdivided into multiple CGGA histologic categories spanning different WHO grades and including both primary and recurrent tumors. This cohort structure highlights the diagnostic and clinical heterogeneity of the CGGA dataset used for cross-cohort BioCoord validation.

**Supplementary Figure S8. Direct analysis of shared TCGA–CGGA genes does not clearly separate CGGA gliomas.** DQC was applied directly to the 657 CGGA tumors with available overall-survival data using the 16,154 genes shared between TCGA and CGGA. Each point represents one tumor. Tumors are shown in a three-dimensional view for display only. Clinical information, including histology, recurrence status, and survival, was not used to build the tumor map. Colors indicate histology added afterward for interpretation: magenta, LGG; blue, GBM. (A–F) Six representative frames show how tumors moved during DQC analysis. Although tumors became more tightly grouped over time, LGG and GBM tumors remained mixed rather than separating into clear clinical groups. These results show that direct analysis of the shared gene set did not recover a clinically useful glioma organization in CGGA, supporting the need for the BioCoord-based validation analysis.

**Fig. S9. Denoising CGGA gene-expression data by feature selection improves visual clustering but does not recover diagnostic or prognostic structure.** To test whether reducing noise in the CGGA gene-expression matrix could improve clustering performance, DQC was applied to a knee-selected reduced gene set of approximately 636 genes. Representative DQC evolution frames are shown: (A) initial frame, (B) intermediate frame, and (C) late frame. Each point represents one tumor. For visualization only, diagnostic labels were applied post hoc: GBM samples are shown in blue, and LGG samples are shown in magenta. These diagnostic annotations were not used for clustering. Compared with the full shared gene-expression matrix, the reduced gene set produced clearer visual contraction and limited local organization during DQC evolution. However, unlike the TCGA discovery analysis, this feature-selected CGGA gene-expression map did not resolve a clear diagnostic structure: LGG and GBM samples remained substantially overlapping, and the resulting clusters did not yield meaningful overall-survival stratification in post hoc analyses. These findings show that feature selection can reduce visual noise, but gene-expression clustering alone was not sufficient to recover clinically meaningful glioma organization in the independent CGGA cohort.

**Supplementary Fig. S10. CGGA tumors organize into clinically recognizable groups in TCGA-derived BioCoord space.** DQC was applied to 693 CGGA tumors using four BioCoords recalculated from CGGA RNA expression with the TCGA-defined BioCoord gene modules. For visualization only, tumors are displayed in the first three principal components of the four-dimensional BioCoord space. Representative DQC frames are shown: (A) initial frame, (B) early frame, (C) intermediate frame, (D–E) late frames, and (F) final frame. Each point represents one tumor. Diagnostic labels, survival, and other clinical annotations were not used during clustering. For post hoc visualization, diagnostic labels were overlaid after clustering: LGG samples are shown in magenta and GBM samples in blue. Cluster labels indicate the approximate locations of the final DQC-defined tumor groups. During DQC evolution, CGGA tumors move from a dispersed pattern into a more organized BioCoord- based tumor map, with two major tumor populations and two smaller separated groups. Intermediate frames best show the relationship among tumor states, whereas later frames show contraction toward final group positions. This analysis shows that TCGA-derived BioCoords preserve clinically recognizable glioma organization in an independent CGGA cohort.

**Supplementary Fig. S11. Overall survival aligns with BioCoord-derived tumor organization in CGGA.** CGGA tumors were projected into TCGA-derived BioCoord space, and overall survival was overlaid after DQC-based tumor groups were defined. Clinical outcome was not used during BioCoord calculation or clustering. (A–C) Three-dimensional visualizations show overall survival distribution across the BioCoord-derived tumor map from different viewing angles. Each point represents one tumor; the vertical axis shows overall survival time. Diagnostic labels are shown post hoc, with LGG samples in magenta and GBM samples in blue. These views show that tumors with different survival outcomes occupy different regions of the BioCoord map, rather than being randomly distributed across the space. (D) Kaplan–Meier analysis comparing BioCoord-derived tumor groups shows clear overall-survival separation, with longer survival in the LGG-enriched BioCoord group and shorter survival in the GBM-enriched BioCoord group. Together, these results show that BioCoord-derived tumor organization in CGGA is associated with overall survival.

**Supplementary Fig. S12. Established clinical and molecular features are associated with overall survival in CGGA but do not fully capture outcome heterogeneity.** To place the BioCoord findings in the context of standard clinical and molecular predictors, overall survival was examined across key CGGA annotations. (A) Median overall survival stratified by WHO grade shows progressively shorter survival from Grade II to Grade IV tumors. (B) Median overall survival stratified by IDH mutation status shows longer survival in IDH-mutant tumors compared with IDH-wildtype tumors. (C) Median overall survival stratified by 1p/19q status shows longer survival in codeleted tumors compared with non-codeleted tumors. (D) Median overall survival stratified by MGMT promoter methylation status shows a modest survival difference between methylated and unmethylated tumors. (E) Kaplan–Meier analysis restricted to WHO Grade III tumors shows clear survival divergence by IDH status, indicating that tumors within the same histologic grade can have different clinical outcomes. Together, these analyses show that established diagnostic and molecular variables are clinically informative, while also highlighting survival heterogeneity that is not fully captured by any single variable alone.

**Supplementary Fig. S13. Survival effects of clinical and molecular features vary across age and treatment contexts in the CGGA cohort.**

To assess whether established prognostic features show consistent survival patterns across clinical contexts, median overall survival (OS, days) was examined after stratification by age and treatment status. (A–C) Median OS by WHO grade is shown for patients aged <40 years, patients aged ≥60 years, and patients aged ≥60 years without treatment, respectively. (D–F) Median OS by IDH mutation status is shown across the same three patient subsets. (G–I) Median OS by MGMT promoter methylation status is shown across the same three patient subsets. (J–L) Median OS by 1p/19q codeletion status is shown across the same three patient subsets. Across these analyses, expected survival trends were generally observed, including shorter survival with higher WHO grade and longer survival with IDH mutation or 1p/19q codeletion. However, the magnitude of these survival differences varied by age and treatment context, indicating that no single clinical or molecular feature fully accounts for outcome heterogeneity in CGGA.

## Supplementary Tables

**Table S1. Mean normalized expression of 20,057 protein-coding genes across DQC- predicted LGG and GBM groups from the initial whole-transcriptome analysis.**

**Table S2. Ranked genes between the Step-1 DQC GBM-enriched and LGG-enriched clusters defining the 554-gene panel.**

**Table S3. IPA-curated annotations for genes ranked by log2 fold change between the Step-1 DQC GBM-enriched and LGG-enriched clusters within the 554-gene panel.**

**Table S4. IPA canonical pathways ranked by Benjamini–Hochberg adjusted p values for genes distinguishing the Step-1 DQC GBM-enriched and LGG-enriched clusters.**

**Table S5. DQC cluster membership and survival summaries for top-k gene panels** Notes: Clusters are derived by DQC without labels. TCGA histology and PFS/OS were applied post hoc for evaluation only. PFS/OS values are reported as mean days.

The rank-ordered gene panels preserve DQC structure and refine LGG prognostic substructure. To assess whether a reduced gene set could preserve diagnostic separation while revealing finer biological structure, genes were ranked by two- sample t-test p-values between the two DQC-defined clusters (used for ordering only), and DQC was evaluated across progressively smaller top-k subsets (140, 100, and 90 genes; **Table S5**).

At 140 genes, DQC resolved three clusters, including two LGG-only groups with distinct survival and one GBM-enriched group. At 100 genes, the geometry refined to four clusters, with three LGG-only groups and one GBM-enriched cluster, indicating improved diagnostic separation. At 90 genes, this four-cluster structure and its associated survival ordering were preserved, supporting a stable low-dimensional representation of the data.

Together, these results indicate that progressive feature reduction denoises the dataset while preserving the DQC manifold, maintaining robust LGG–GBM separation, and exposing reproducible prognostic substructure within LGG. Performance deteriorated below ∼90 genes, suggesting a stable “sweet spot” near 90–100 genes.

**Table S6. Clinical and molecular characteristics of the CGGA validation cohort**

**Table S7. Clinical and molecular annotations of the CGGA validation cohort by BioCoord-derived tumor group.xlsx**

## References

1. Galbraith K, Snuderl M. Molecular Pathology of Gliomas. (2021) Surg Pathol Clin 14(3):379-386. doi: 10.1016/j.path.2021.05.003.

2. Nafe R, Porto L, Samp PF, et al. (2023) Adult-type and Pediatric-type Diffuse Gliomas : What the Neuroradiologist Should Know. Clin Neuroradiol 33(3):611–624. doi: 10.1007/s00062-023-01277-z.

3. Touat M, Li YY, Boynton AN, et al. (2020) Mechanisms and therapeutic implications of hypermutation in gliomas. Nature. 580(7804):517–523. doi: 10.1038/s41586-020-2209-9. Epub 2020 Apr 15. PMID: 32322066; PMCID: PMC8235024.

4. Ma S, Guo Z, Wang B, et al. (2022) A Computational Framework to Identify Biomarkers for Glioma Recurrence and Potential Drugs Targeting Them. Front Genet. 12:832627. doi: 10.3389/fgene.2021.832627.

5. Louis DN, Perry A, Wesseling P, et al. (2021) The 2021 WHO Classification of Tumors of the Central Nervous System: a summary. Neuro Oncol. 2021; 23(8):1231-1251. doi: 10.1093/neuonc/noab106. PMID: 34185076; PMCID: PMC8328013.

6. Ispirjan M, Marx S, Freund E, et al. (2024) Markers of tumor-associated macrophages and microglia exhibit high intratumoral heterogeneity in human glioblastoma tissue. Oncoimmunology 13(1):2425124. doi: 10.1080/2162402X.2024.2425124.

7. Inano, R., Oishi, N., Kunieda, T. et al. (2016) Visualization of heterogeneity and regional grading of gliomas by multiple features using magnetic resonance-based clustered images. Sci Rep 6, 30344. 10.1038/srep30344

8. Linninger A, Hartung GA, Liu BP, et al. (2018) Modeling the diffusion of D-2- hydroxyglutarate from IDH1 mutant gliomas in the central nervous system. Neuro Oncol 20(9):1197–1206. doi: 10.1093/neuonc/noy051.

9. Berger A, Tzarfati, GG, Serafimova M, et al. (2022) Clinical and prognostic implications of rim restriction following glioma surgery. Sci Rep 12, 12874. 10.1038/s41598-022-16717-y

10. Zhao WJ, Ou GY, Lin WW. (2021) Integrative Analysis of Neuregulin Family Members- Related Tumor Microenvironment for Predicting the Prognosis in Gliomas. Front Immunol. 12:682415. doi: 10.3389/fimmu.2021.682415.

11. Miniere HJM, Hormuth DA 2nd, Lima EABF, et al. (2025) A data assimilation framework for predicting the spatiotemporal response of high-grade gliomas to chemoradiation. BMC Cancer 25(1):1239. doi: 10.1186/s12885-025-14557-3.

12. Bähr O, Herrlinger U, Weller M, Steinbach JP. (2009) Very late relapses in glioblastoma long-term survivors. J Neurol. 256(10):1756–8. doi: 10.1007/s00415-009-5167-6.

13. Stupp R, Mason WP, van den Bent MJ, et al. (2005) Radiotherapy plus concomitant and adjuvant temozolomide for glioblastoma. N Engl J Med. 352(10):987–96. doi: 10.1056/NEJMoa043330. PMID: 15758009.

14. Ceccarelli M, Barthel FP, Malta TM, et al. (2016) Molecular Profiling Reveals Biologically Discrete Subsets and Pathways of Progression in Diffuse Glioma. Cell 164(3):550–63. doi: 10.1016/j.cell.2015.12.028.

15. Cancer Genome Atlas Research Network; Brat DJ, Verhaak RG, et al. (2015) Comprehensive, Integrative Genomic Analysis of Diffuse Lower-Grade Gliomas. N Engl J Med. 372(26):2481-98. doi: 10.1056/NEJMoa1402121.

16. Komori T. (2022) Grading of adult diffuse gliomas according to the 2021 WHO Classification of Tumors of the Central Nervous System. Lab Invest. 102(2):126–133. doi: 10.1038/s41374-021-00667-6.

17. Neftel C, Laffy J, Filbin MG, et al. (2019) An Integrative Model of Cellular States, Plasticity, and Genetics for Glioblastoma. Cell. 178(4):835–849.e21. doi: 10.1016/j.cell.2019.06.024.

18. Wang Q, Hu B, Hu X, et al. (2017) Tumor Evolution of Glioma-Intrinsic Gene Expression Subtypes Associates with Immunological Changes in the Microenvironment. Cancer Cell. 2017 Jul 10;32(1):42-56.e6. doi: 10.1016/j.ccell.2017.06.003.

19. The Cancer Genome Atlas Research Network., Weinstein, J., Collisson, E., et al. (2013) The Cancer Genome Atlas Pan-Cancer analysis project. Nat Genet 45: 1113–1120. 10.1038/ng.2764

20. GTEx Consortium. Human genomics. (2015) The Genotype-Tissue Expression (GTEx) pilot analysis: multitissue gene regulation in humans. Science. 348(6235):648–60. doi: 10.1126/science.1262110.

21. Mailman MD, Feolo M, Jin Y, et al. (2007) The NCBI dbGaP database of genotypes and phenotypes. Nat Genet. 39(10):1181–6. doi: 10.1038/ng1007-1181. PMID: 17898773; PMCID: PMC2031016.

22. Zhao Z, Zhang KN, Wang Q, et al. (2021) Chinese Glioma Genome Atlas (CGGA): A Comprehensive Resource with Functional Genomic Data from Chinese Glioma Patients. Genomics Proteomics Bioinformatics.19(1):1–12. doi: 10.1016/j.gpb.2020.10.005. Epub 2021 Mar 2. PMID: 33662628; PMCID: PMC8498921.

23. Cui W, Xue H, Wei L, et al. (2021) High heterogeneity undermines generalization of differential expression results in RNA-Seq analysis. Hum Genomics. 15(1):7. doi:10.1186/s40246-021-00308-5.

24. Greenwald AC, Darnell NG, Hoefflin R, et al. (2024) Integrative spatial analysis reveals a multi-layered organization of glioblastoma. Cell. 187(10):2485–2501.e26. doi: 10.1016/j.cell.2024.03.029. Epub 2024 Apr 22. PMID: 38653236; PMCID: PMC11088502.

25. Bulik-Sullivan B, Finucane HK, Anttila V, et al. (2015) An atlas of genetic correlations across human diseases and traits. Nat Genet. 47(11):1236–41. doi: 10.1038/ng.3406. Epub 2015 Sep 28. PMID: 26414676; PMCID: PMC4797329.

26. Echle A, Rindtorff NT, Brinker TJ et al. (2021) Deep learning in cancer pathology: a new generation of clinical biomarkers. Br J Cancer 124: 686–696. 10.1038/s41416-020-01122-x

27. Yu Y, Mai Y, Zheng Y, et al. (2024) Assessing and mitigating batch effects in large-scale omics studies. Genome Biol 25: 254. 10.1186/s13059-024-03401-9.

28. Roth A, Khattra J, Yap D, et al. (2014) PyClone: statistical inference of clonal population structure in cancer. Nat Methods 11: 396–398. 10.1038/nmeth.2883.

29. Wang Z, Cao S, Morris JS, et al. (2018) Transcriptome Deconvolution of Heterogeneous Tumor Samples with Immune Infiltration. iScience 9:451–460. doi: 10.1016/j.isci.2018.10.028.

30. Leek JT, Scharpf RB, Bravo HC et al. (2010) Tackling the widespread and critical impact of batch effects in high-throughput data. Nat Rev Genet. 11(10):733–9. doi: 10.1038/nrg2825.

31. Jolliffe IT, Cadima J. (2016) Principal component analysis: a review and recent developments. Philos Trans A Math Phys Eng Sci. 13;374(2065):20150202.

32. Kobak D, Berens P. (2019) The art of using t-SNE for single-cell transcriptomics. Nat Commun. 10(1):5416.

33. Chaudhry M, Shafi I, Mahnoor M, et al. (2023). A Systematic Literature Review on Identifying Patterns Using Unsupervised Clustering Algorithms: A Data Mining Perspective. Symmetry 15(9): 1679. 10.3390/sym15091679.

34. Wani AA (2025). Comprehensive review of dimensionality reduction algorithms: challenges, limitations, and innovative solutions. PeerJ Computer Science 11: e3025. 10.7717/peerj-cs.3025

35. Wang Y, Huang H, Rudin C, Shaposhnik Y. (2021) Understanding how dimension reduction tools work: An empirical approach to deciphering t-SNE, UMAP, TriMap, and PaCMAP for data visualization. J Machine Learning Res 22:1–73. 10.5555/3546258.3546459.

36. Weinstein M, Horn D. (2009) Dynamic quantum clustering: A method for visual exploration of structures in data. Phys Rev E Stat Nonlin Soft Matter Phys. 80(6 Pt 2): Article 066117. 10.1103/PhysRevE.80.066117.

37. Weinstein M, Meirer F, et al. (2013) Analyzing big data with dynamic quantum clustering. arXiv preprint arXiv:1310.2700. 10.48550/arXiv.1310.2700.

38. Roche KE, Weinstein M, Dunwoodie LJ, et al. (2018) Sorting Five Human Tumor Types Reveals Specific Biomarkers and Background Classification Genes. Sci Rep 8: 8180. 10.1038/s41598-018-26310-x.

39. Wang Z, Gerstein M, Snyder M. (2009) RNA-Seq: a revolutionary tool for transcriptomics. Nat Rev Genet 10:57–63.

40. Coifman RR, Lafon S, Lee AB, et al. (2005) Geometric diffusions as a tool for harmonic analysis and structure definition of data: diffusion maps. Proc Natl Acad Sci U S A: 102(21):7426–31.

41. Reimand J, Isserlin R, Voisin V, et al. (2019) Pathway enrichment analysis and visualization of omics data using g:Profiler, GSEA, Cytoscape and EnrichmentMap. Nat Protoc. 14(2):482–517. doi: 10.1038/s41596-018-0103-9.

42. Tomfohr J, Lu J, Kepler TB. (2005) Pathway level analysis of gene expression using singular value decomposition. BMC Bioinformatics. 6:225. doi:10.1186/1471-2105-6-225.

43. Albrecht LJ, Höwner A, Griewank K, et al. (2022) Circulating cell-free messenger RNA enables non-invasive pan-tumour monitoring of melanoma therapy independent of the mutational genotype. Clin Transl Med. 12(11):e1090. doi: 10.1002/ctm2.1090.

44. Kaufman S, Rosset S, Perlich C, Stitelman O. (2012) Leakage in data mining: Formulation, detection, and avoidance. ACM Transactions on Knowledge from Data (TKDD): 6(4):1–21. 10.1145/2382577.2382579

45. Sivakumar M, Parthasarathy S, Padmapriya T. (2024) Trade-off between training and testing ratio in machine learning for medical image processing. PeerJ Comput Sci. 10:e2245. doi: 10.7717/peerj-cs.2245.

46. Cawley GC, Talbot NLC (2010) On over-fitting in model selection and subsequent selection bias in performance evaluation. The Journal of Machine Learning Research 11:2079–2107.

47. Jing L, Vincent P, LeCun Y, Tian Y (2022) Understanding dimensional collapse in contrastive self-supervised learning. Proceedings of the 10th International Conference on Learning Representations (ICLR).

48. Krämer A, Green J, Pollard Jr J, Tugendreich S. (2014) Causal analysis approaches in Ingenuity Pathway Analysis. Bioinformatics 30(4):523–30.

49. Benjamini Y & Hochberg Y. (1995). Controlling the false discovery rate: A practical and powerful approach to multiple testing. Journal of the Royal Statistical Society: Series B (Methodological), 57(1), 289–300.

50. Lönnstedt I, Speed T (2002) Replicated microarray data. Statistica Sinica 12(1) A Special Issue on Bioinformatics:31-46.

51. Liberzon A, Birger C, Thorvaldsdóttir H, et al. (2015) The Molecular Signatures Database (MSigDB) hallmark gene set collection. Cell Syst. 1(6):417–425. doi: 10.1016/j.cels.2015.12.004.

52. Brunet J-P. Tamayo P, Golub TR, Mesirov JP. (2004) Metagenes and molecular pattern discovery using matrix factorization. Proc Natl Acad Sci U.S.A. 101(12):4164–9. doi:10.1073/pnas.0308531101

53. Zito A, Escribà Montagut X, et al. (2025) PLAID: ultrafast single-sample gene set enrichment scoring. Bioinformatics. 41(12):btaf621. doi: 10.1093/bioinformatics/btaf621. PMID: 41223139; PMCID: PMC12694415.

54. McDermaid A, Monier B, Zhao J, Liu B, Ma Q. (2019) Interpretation of differential gene expression results of RNA-seq data: review and integration. Brief Bioinform. 20(6):2044– 2054. doi: 10.1093/bib/bby067

55. Satopää V, Albrecht J, Irwin D, Raghavan B. (2011). Finding a "Kneedle" in a Haystack: Detecting Knee Points in System Behavior. In 2011 31st International Conference on Distributed Computing Systems Workshops (pp. 166-171). IEEE. 10.1109/ICDCSW.2011.20

56. Nicholson JG, Fine HA. (2021) Diffuse Glioma Heterogeneity and Its Therapeutic Implications. Cancer Discov. 11(3):575–590. doi: 10.1158/2159-8290.CD-20-1474. Epub 2021 Feb 8. PMID: 33558264.

57. Antonelli M, Poliani PL. (2022) **Adult type diffuse gliomas in the new** 2021 **WHO Classification**. Pathologica. 114(6):397–409. doi: 10.32074/1591-951X-823. PMID: 36534419; PMCID: PMC9763975.

58. Varn FS, Johnson KC, Martinek J, et al. (2022) Glioma progression is shaped by genetic evolution and microenvironment interactions. Cell. 185(12):2184–2199.e16. doi: 10.1016/j.cell.2022.04.038. Epub 2022 May 31. PMID: 35649412; PMCID: PMC9189056.

59. Phillips HS, Kharbanda S, Chen R, et al. (2006) Molecular subclasses of high-grade glioma predict prognosis, delineate a pattern of disease progression, and resemble stages in neurogenesis. Cancer Cell. 9(3):157–73. doi: 10.1016/j.ccr.2006.02.019. PMID: 16530701.

60. Johnson KC, Spitzer A, Varn FS, et al. (2026) Acquired genetic and cell-state changes in IDH-mutant glioma progression. Nature. 655(8124):1048–1059. doi: 10.1038/s41586-026-10612-6. Epub 2026 Jun 3. PMID: 42236943; PMCID: PMC13391360.

61. Venteicher AS, Tirosh I, Hebert C, et al. (2017) Decoupling genetics, lineages, and microenvironment in IDH-mutant gliomas by single-cell RNA-seq. Science. 355(6332):eaai8478. doi: 10.1126/science.aai8478. PMID: 28360267; PMCID: PMC5519096.

62. Wu J, Gonzalez Castro LN, et al. (2025) Evolving cell states and oncogenic drivers during the progression of IDH-mutant gliomas. Nat Cancer. 6(1):145–157. doi: 10.1038/s43018-024-00865-3. Epub 2024 Nov 21. PMID: 39572850.

63. Le Boiteux E, Court F, Guichet PO, et al. (2021) Widespread overexpression from the four DNA hypermethylated HOX clusters in aggressive (IDHwt) glioma is associated with H3K27me3 depletion and alternative promoter usage. Mol Oncol. 15(8):1995–2010. doi: 10.1002/1878-0261.12944. Epub 2021 May 2. PMID: 33720519; PMCID: PMC8334257.

64. Hegi ME, Diserens AC, Gorlia T, et al. (2005) MGMT gene silencing and benefit from temozolomide in glioblastoma. N Engl J Med. 352(10):997–1003. doi: 10.1056/NEJMoa043331. PMID: 15758010.

65. Bhat KPL, Balasubramaniyan V, Vaillant B, et al. (2013) Mesenchymal differentiation mediated by NF-κB promotes radiation resistance in glioblastoma. Cancer Cell. 24(3):331–46. doi: 10.1016/j.ccr.2013.08.001. Epub 2013 Aug 29. PMID: 23993863; PMCID: PMC3817560.

66. Spitzer A, Johnson KC, Nomura M, et al. (2025) Deciphering the longitudinal trajectories of glioblastoma ecosystems by integrative single-cell genomics. Nat Genet. 57(5):1168–1178. doi: 10.1038/s41588-025-02168-4. Epub 2025 May 9. PMID: 40346362; PMCID: PMC12081298.

67. Murat A, Migliavacca E, Gorlia T, et al. (2008) Stem cell-related "self-renewal" signature and high epidermal growth factor receptor expression associated with resistance to concomitant chemoradiotherapy in glioblastoma. J Clin Oncol. 26(18):3015–24. doi: 10.1200/JCO.2007.15.7164. PMID: 18565887.

68. Tirosh I, Venteicher AS, Hebert C, et al. (2016) Single-cell RNA-seq supports a developmental hierarchy in human oligodendroglioma. Nature. 539(7628):309–313. doi: 10.1038/nature20123. Epub 2016 Nov 2. PMID: 27806376; PMCID: PMC5465819.

69. Barthel FP, Johnson KC, Varn FS, et al. (2019) Longitudinal molecular trajectories of diffuse glioma in adults. Nature. 576(7785):112–120. doi: 10.1038/s41586-019-1775-1. Epub 2019 Nov 20. PMID: 31748746; PMCID: PMC6897368.

70. Gonçalves CS, Le Boiteux E, Arnaud P, Costa BM. (2020) HOX gene cluster (de)regulation in brain: from neurodevelopment to malignant glial tumours. Cell Mol Life Sci. 77(19):3797–3821. doi: 10.1007/s00018-020-03508-9. Epub 2020 Apr 1. PMID: 32239260; PMCID: PMC11105007.

71. He ZC, Liu Q, Yang KD, et al. (2022) HOXA5 is amplified in glioblastoma stem cells and promotes tumor progression by transcriptionally activating PTPRZ1. Cancer Lett. 533:215605. doi: 10.1016/j.canlet.2022.215605. Epub 2022 Feb 24. PMID: 35219772.

72. Patel AP, Tirosh I, Trombetta JJ, et al. (2014) Single-cell RNA-seq highlights intratumoral heterogeneity in primary glioblastoma. Science. 344(6190):1396–401. doi: 10.1126/science.1254257. Epub 2014 Jun 12. PMID: 24925914; PMCID: PMC4123637.

73. Verhaak RG, Hoadley KA, Purdom E, et al. (2010) Integrated genomic analysis identifies clinically relevant subtypes of glioblastoma characterized by abnormalities in PDGFRA, IDH1, EGFR, and NF1. Cancer Cell.17(1):98-110. doi: 10.1016/j.ccr.2009.12.020. PMID: 20129251; PMCID: PMC2818769.

74. Carro MS, Lim WK, Alvarez MJ, et al. (2026) Longitudinal changes in DNA methylation in IDH-mutant glioma fuel disease progression through altered cell state differentiation. Nat Genet. 58(7):1651–1660. doi: 10.1038/s41588-026-02642-7. Epub 2026 Jun 22. PMID: 42332268; PMCID: PMC13364636.

76. Domínguez Conde C, Xu C, Jarvis LB, et al. (2022) Cross-tissue immune cell analysis reveals tissue-specific features in humans. Science. 376(6594):eabl5197. doi: 10.1126/science.abl5197. Epub 2022 May 13. PMID: 35549406; PMCID: PMC7612735.

77. Shi Q, Tang F, Chen Y, Zhang Z. (2026) Multicellular ecosystems: Linking cellular diversity to tissue function and disease. Trends Cell Biol. S0962–8924(26)00104-2. doi: 10.1016/j.tcb.2026.06.005. Epub ahead of print. PMID: 42342504.

78. Galindez G, Sadegh S, Baumbach J, Kacprowski T, List M. (2022) Network-based approaches for modeling disease regulation and progression. Comput Struct Biotechnol J. 21:780–795. doi: 10.1016/j.csbj.2022.12.022. PMID: 36698974; PMCID: PMC9841310.

79. Mathur R, Wang Q, Schupp PG, et al. (2024) Glioblastoma evolution and heterogeneity from a 3D whole-tumor perspective. Cell. 187(2):446–463.e16. doi: 10.1016/j.cell.2023.12.013. PMID: 38242087; PMCID: PMC10832360.

80. Puchalski RB, Shah N, Miller J, et al. (2018) An anatomic transcriptional atlas of human glioblastoma. Science. 360(6389):660–663. doi: 10.1126/science.aaf2666. PMID: 29748285; PMCID: PMC6414061.

81. Sottoriva A, Spiteri I, Piccirillo SG, et al. (2013) Intratumor heterogeneity in human glioblastoma reflects cancer evolutionary dynamics. Proc Natl Acad Sci U S A. 110(10):4009–14. doi: 10.1073/pnas.1219747110. Epub 2013 Feb 14. PMID: 23412337; PMCID: PMC3593922.

## Supplemental references

1. Weinstein M, Horn D. Dynamic quantum clustering: A method for visual exploration of structures in data. Phys Rev E Stat Nonlin Soft Matter Phys. 2009;80(6 Pt 2): Article 066117. 10.1103/PhysRevE.80.066117.

2. Weinstein M., Meirer F., et al. (2013) Analyzing big data with dynamic quantum clustering. arXiv preprint arXiv:1310.2700. 10.48550/arXiv.1310.2700.

