## Supplemental Document for "Dynamic Quantum Clustering of Glioma RNA-seq Reveals Interpretable Tumor States Linked to Diagnosis and Survival"

**Supplementary Note S1-1. Intuitive description of Dynamic Quantum Clustering (DQC)**

The TCGA glioma analysis revealed that extended structures emerging during DQC evolution, even when applied to noisy high-dimensional expression data, correspond to clinically meaningful patterns such as progression-free survival (PFS) and overall survival (OS). To help readers understand why this occurs, we provide an intuitive explanation of the DQC procedure using a simple two-dimensional example. A full mathematical description of the algorithm is available in the original publications (Weinstein & Horn, Phys. Rev. E 80, 066117; Horn & Gottlieb, arXiv:1310.2700).

The explanation below avoids mathematical formalism and instead illustrates the geometric intuition underlying the algorithm.

---

**Supplementary Note S1-2. Model dataset used for illustration**

To illustrate how DQC reveals hidden structure in complex data, we generated a two-dimensional dataset intended to mimic simplified RNA-seq measurements for two genes. The dataset contains four components:

1. **Three compact Gaussian clusters**, each representing a group of tumors with similar expression values. Differences among points within each cluster arise only from statistical variability.
2. **One extended distribution**, constructed by concatenating many overlapping Gaussian distributions arranged along a vertical trajectory. This extended structure represents a set of tumors whose gene expression changes smoothly along an underlying biological axis (for example, disease progression).

The combined dataset therefore contains both compact clusters and continuous structure, similar to patterns commonly observed in transcriptomic data.

**Figure S1** shows the simulated dataset.

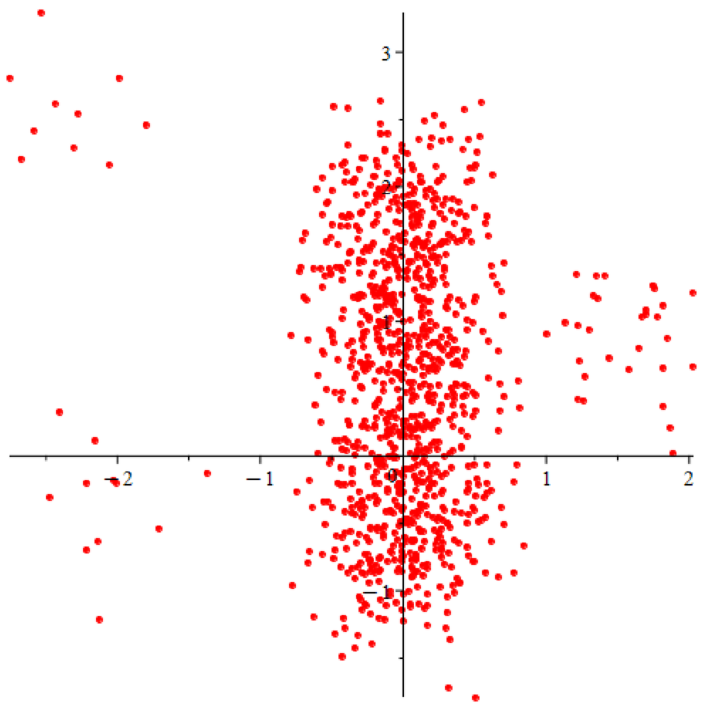


**Figure S1. Simulated dataset used to illustrate DQC behavior.**

Two-dimensional model dataset composed of three compact Gaussian clusters and one extended distribution generated from overlapping Gaussian components arranged along a vertical trajectory. The simulated data are used to illustrate how DQC distinguishes compact groupings from continuous structure.

---

**Supplementary Note S1-3. Construction of the DQC potential function**

DQC begins by constructing a potential function from the data.
This function can be understood as a smooth energy-like landscape defined across the data space.

Importantly:

- Regions containing many nearby samples correspond to lower values of the potential function.
- Regions with few samples correspond to higher values.

In intuitive terms, the potential surface highlights areas where samples accumulate.

To visualize this surface, the potential is plotted as a three-dimensional surface above the two-dimensional data plane. The third dimension does not represent an additional data variable; it simply displays the value of the potential function. For visualization, the hyperbolic tangent of the potential (tanh(V)) is shown to suppress large amplitudes away from the data and emphasize the topology near dense regions.

**Figure S2** illustrates this potential surface together with the original data points.

In this representation:

- Compact clusters appear as circular depressions in the landscape.
- Sparse regions may contain multiple small local minima reflecting sampling noise.
- Extended data structures generate elongated valleys in the potential surface.

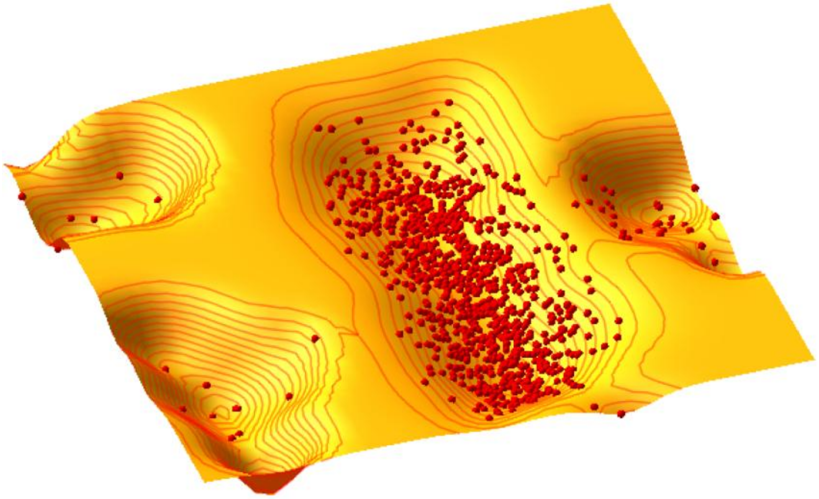

**Figure S2. Potential function generated from the model dataset.**

The DQC potential function is visualized as a three-dimensional surface above the two-dimensional data plane. Lower regions correspond to areas containing higher densities of samples. Data points are shown in red.

This example illustrates how DQC can reveal hidden low-dimensional structure embedded within complex datasets. The full animation can be seen in this file (DQC_Evolution_Dynamics).

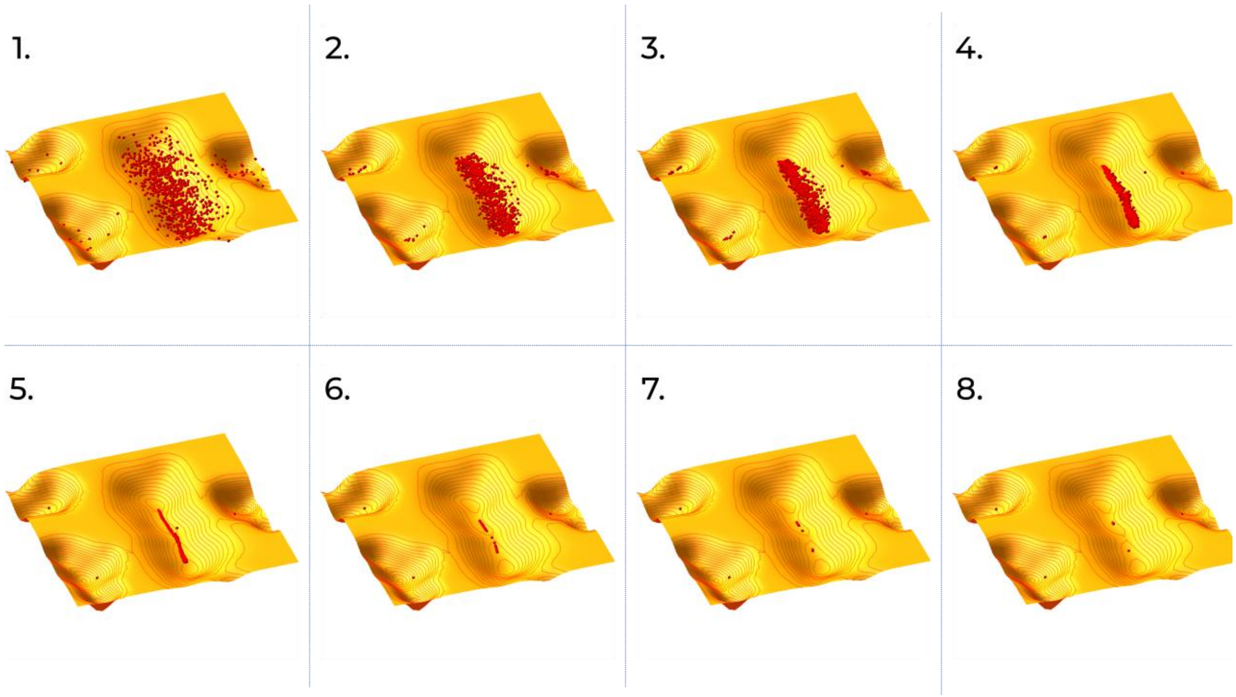


**Figure S3. Evolution of the model dataset during DQC.**

Panels S3-1 to S3-8) Selected frames from the DQC animation showing how samples move across the potential surface over the course of the algorithm . Compact clusters converge to stable basins, whereas the extended distribution contracts into a lower-dimensional structure that reflects the dominant organization of the data.

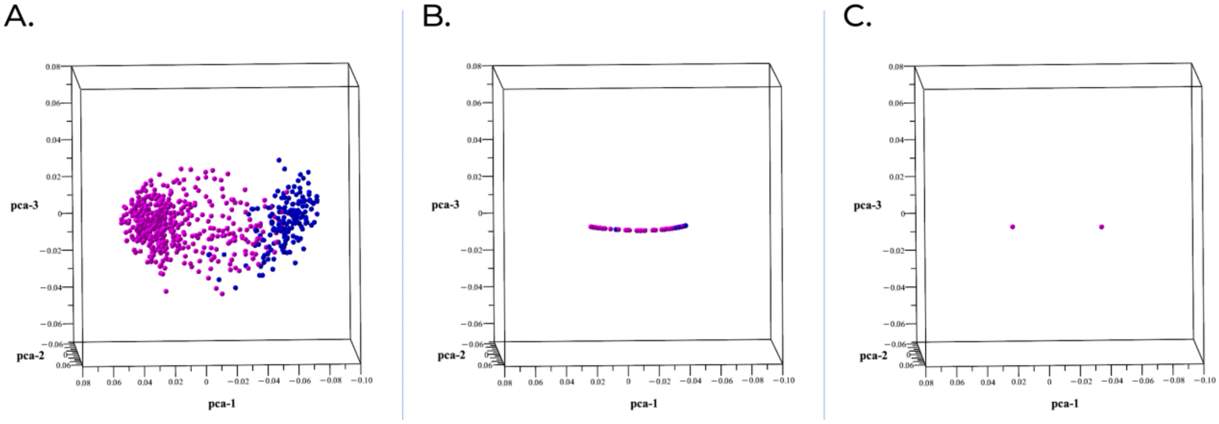


**Figure S4. DQC evolution of the 554-gene panel from diffuse cloud to two final clusters.**

Representative frames from Dynamic Quantum Clustering (DQC) applied to the 692×554 expression matrix, displayed in the first three principal components for visualization only. Left, frame 1: the initial diffuse cloud shows separation of the two dominant sample groupings. Middle, frame 11: the cloud contracts onto a lower-dimensional manifold while preserving the global LGG–GBM organization. Right, final frame: the data resolve into two compact endpoint clusters corresponding to the LGG-enriched and GBM-enriched states identified in the main analysis. Histology labels were overlaid only after clustering for post hoc evaluation.

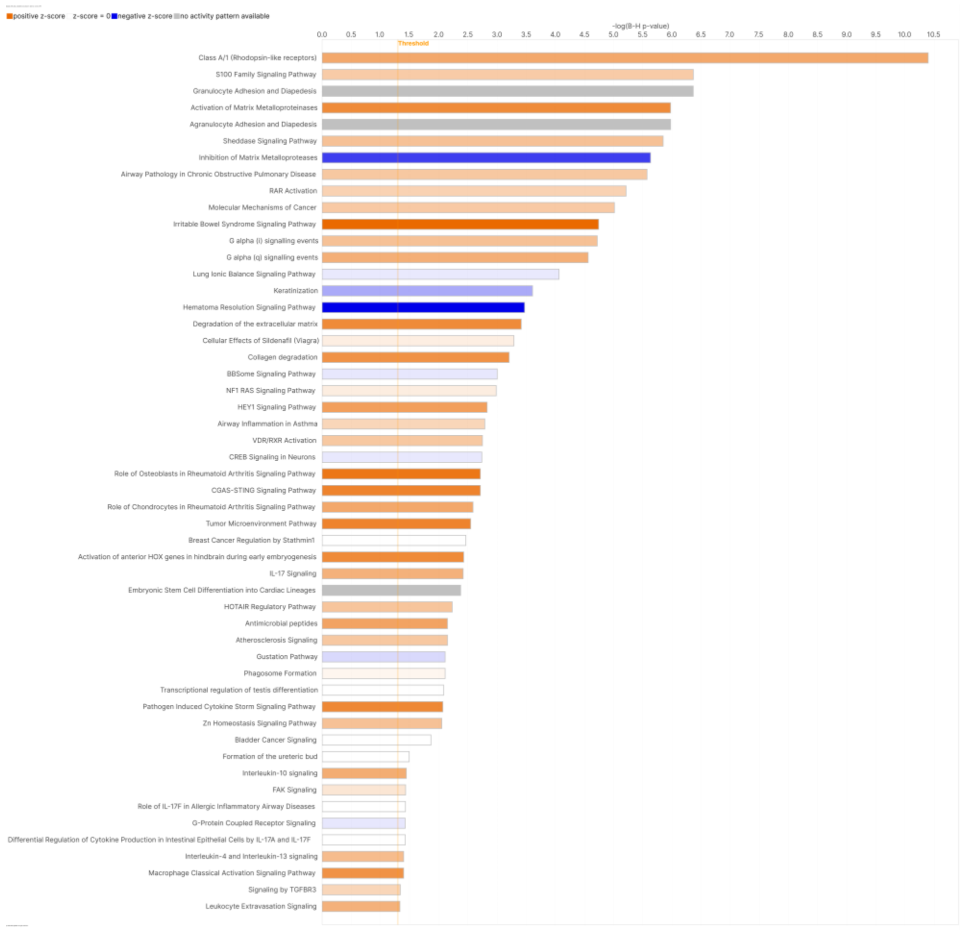


**Figure S5. Pathway enrichment highlights ECM remodeling, inflammation, and invasion programs distinguishing DQC-defined clusters.**

Ingenuity Pathway Analysis (IPA) of differentially expressed genes between the DQC-defined GBM-enriched and LGG-enriched clusters (FDR < 0.05) identified significant enrichment of extracellular matrix (ECM) organization and degradation pathways, including matrix metalloprotease activity and collagen-related processes. Additional enriched pathways include leukocyte adhesion and diapedesis, vascular transmigration, PI3K-associated motility, and pro-inflammatory signaling (e.g., IL-17, NF-κB, and PI3-family pathways). Pathways are ranked by Benjamini–Hochberg–adjusted p values. These results provide biological context for the unsupervised DQC separation, indicating that the GBM-enriched cluster is associated with ECM remodeling, invasion, and inflammatory signaling, whereas the LGG-enriched cluster retains more neuronal and developmental features.

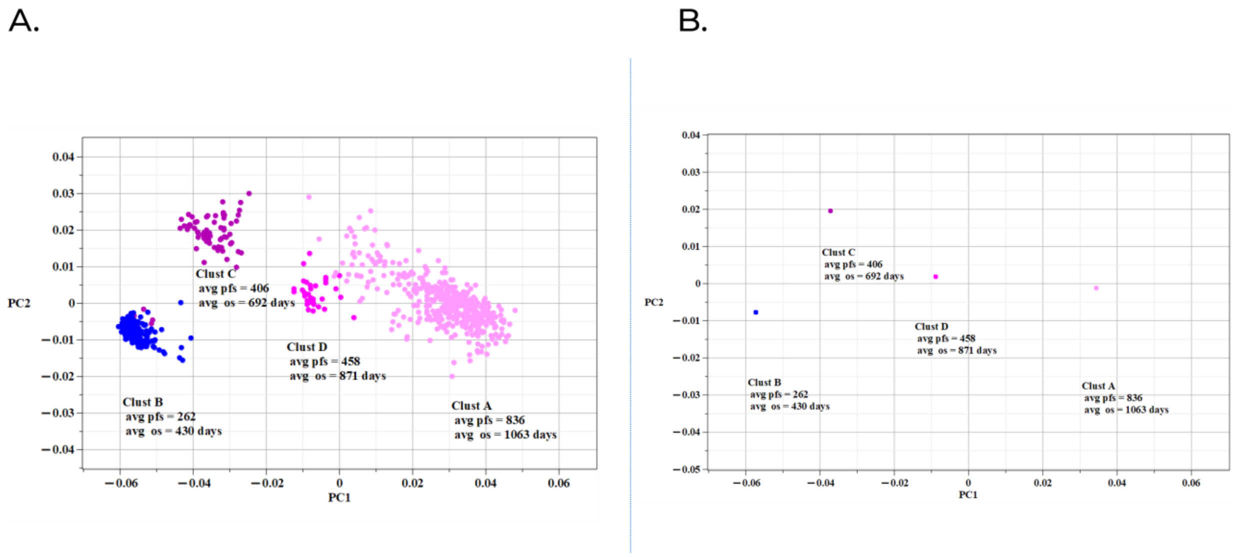


**Figure S6. Compression of the 90-gene DQC structure into biological coordinates for risk stratification**

DQC-based clustering using the 90-gene signature. An unsupervised reduction of the 554-gene set, driven by rank-ordering of genes by between-cluster contrast, yielded a 90-gene panel. Directed Quantum Clustering (DQC)resolved tumors into four clusters (A–D) without the use of clinical labels. While Cluster B is GBM-enriched, notably shows five LGG samples that cluster biologically with the GBM group, suggesting an aggressive molecular phenotype. The clusters occupy an elongated trajectory (A→D→C→B) consistent with a dominant mesenchymal/invasion-associated gradient.

**Supplementary Table S5. DQC cluster composition by TCGA histology and mean survival for the 140-, 100-, and 90-gene panels**

Notes: Clusters are derived by DQC without labels. TCGA histology and PFS/OS were applied post hoc for evaluation only. PFS/OS values are reported as mean days.

**Table S5-1-1. TCGA histology within the three groups identified by DQC using 140 genes**

| | Cluster 140A | Cluster 140B | Cluster 140C |
|---|---|---|---|
| True LGG | 450 | 9 | 65 |
| True GBM | 0 | 168 | 0 |

**Table S5-1-2. Mean progression-free and overall survival in the 140-gene DQC groups**

| Cluster | Avg. PFS | Avg. OS |
|---|---|---|
| Cluster 140A | 811.01 | 1053.23 |
| Cluster 140B | 275.26 | 444.76 |
| Cluster 140C | 389.89 | 659.86 |

**Table S5-2-1. TCGA histology within the four groups identified by DQC using 100 genes**

| | Cluster 100A | Cluster 100B | Cluster 100C | Cluster 100D |
|---|---|---|---|---|
| TCGA LGG | 421 | 5 | 65 | 33 |
| TCGA GBM | 0 | 168 | 0 | 0 |

**Table S5-2-2. Mean progression-free and overall survival in the 100-gene DQC groups**

| Cluster | Avg. PFS | Avg. OS |
|---|---|---|
| Cluster 100A | 835.49 | 1061.96 |
| Cluster 100B | 262.65 | 430.05 |
| Cluster 100C | 397.34 | 602.26 |
| Cluster 100D | 448.42 | 1043.55 |

**Table S5-3-1. TCGA histology within the four groups identified by DQC using 90 genes**

| | Cluster 90A | Cluster 90B | Cluster 90C | Cluster 90D |
|---|---|---|---|---|
| TCGA LGG | 421 | 5 | 65 | 33 |
| TCGA GBM | 0 | 168 | 0 | 0 |

**Table S5-3-2. Mean progression-free and overall survival in the 90-gene DQC groups**

| Cluster | Avg. PFS | Avg. OS |
|---|---|---|
| Cluster 90A | 836.31 | 1062.83 |
| Cluster 90B | 262.65 | 430.05 |
| Cluster 90C | 405.85 | 691.82 |
| Cluster 90D | 457.94 | 871.12 |

**Table S5. DQC cluster composition by TCGA histology and mean survival for the 140-, 100-, and 90-gene panels**

Genes from the 554-gene panel were ranked by two-sample *t*-test *P* values between the two DQC-defined groups. The test was used only to order the genes. DQC was then repeated using the top 140, 100, and 90 genes. TCGA histology and survival information were not used to form the clusters but were added afterward to evaluate their clinical relevance. Using 140 genes, DQC identified three clusters: two LGG-only groups with different mean survival and one GBM-enriched group containing all 168 GBMs and nine LGGs. Using 100 genes, DQC identified four clusters: three LGG-only groups and one GBM-enriched group containing all 168 GBMs and five LGGs. The 90-gene panel preserved the same four-cluster histologic composition and the survival order A → D → C → B. Together, these results show that reducing the gene set to 90 genes preserved the main LGG–GBM separation while revealing LGG groups with distinct clinical outcomes. Among the panels presented in this table, the 90-gene panel was the smallest that preserved this four-group pattern. When fewer than 90 genes were used, the four-cluster pattern was no longer consistently preserved (data not shown).

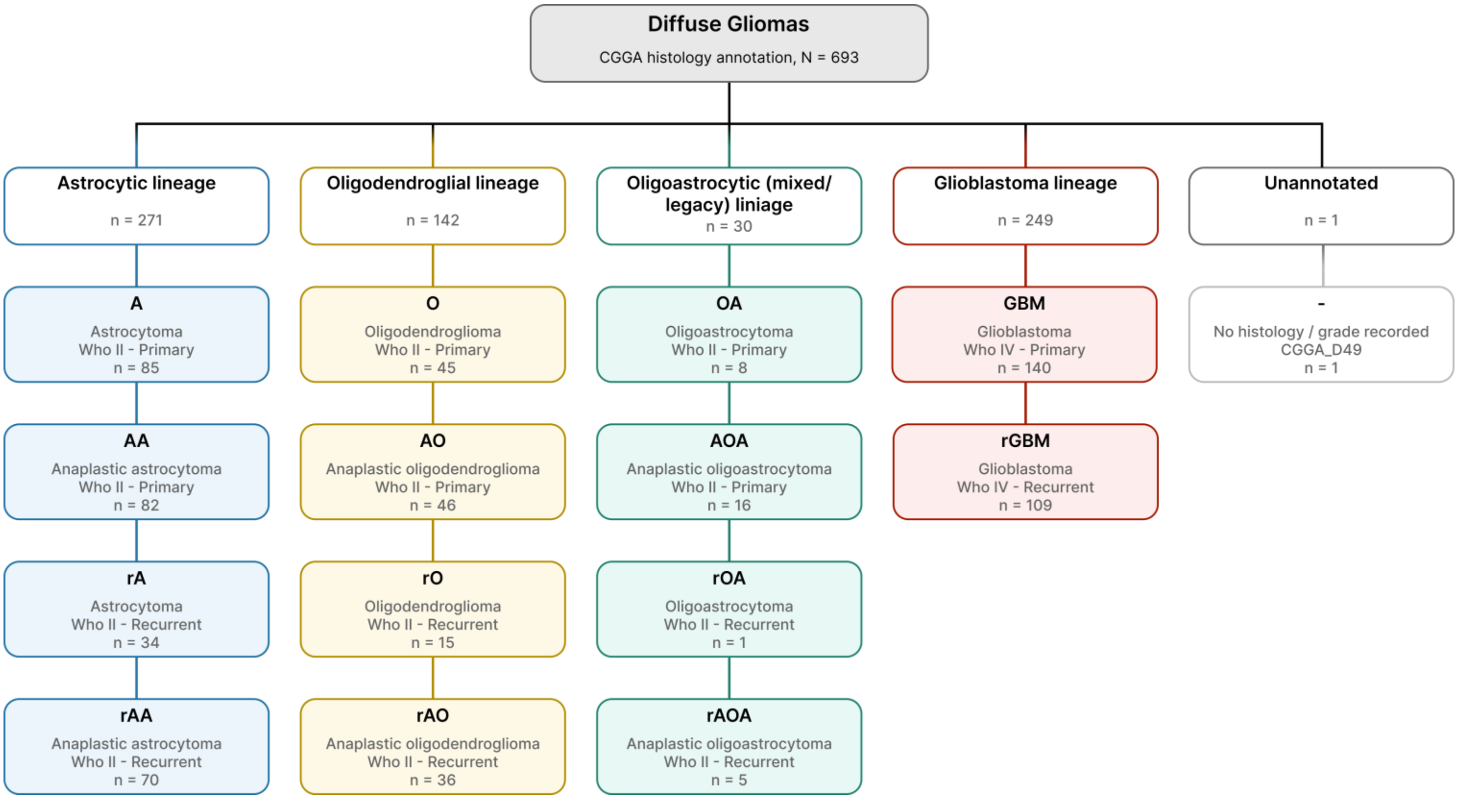


**Supplementary Figure S7. Clinical and histologic heterogeneity of the CGGA validation cohort.**

CGGA clinical metadata were used to summarize the histologic composition of the independent validation cohort. The cohort included 693 diffuse gliomas spanning astrocytic lineage, oligodendroglial lineage, mixed/legacy oligoastrocytic lineage, and glioblastoma lineage, with one tumor lacking histology/grade annotation. Within each lineage, tumors were subdivided into multiple CGGA histologic categories spanning different WHO grades and including both primary and recurrent tumors. This cohort structure highlights the diagnostic and clinical heterogeneity of the CGGA dataset used for cross-cohort BioCoord validation.

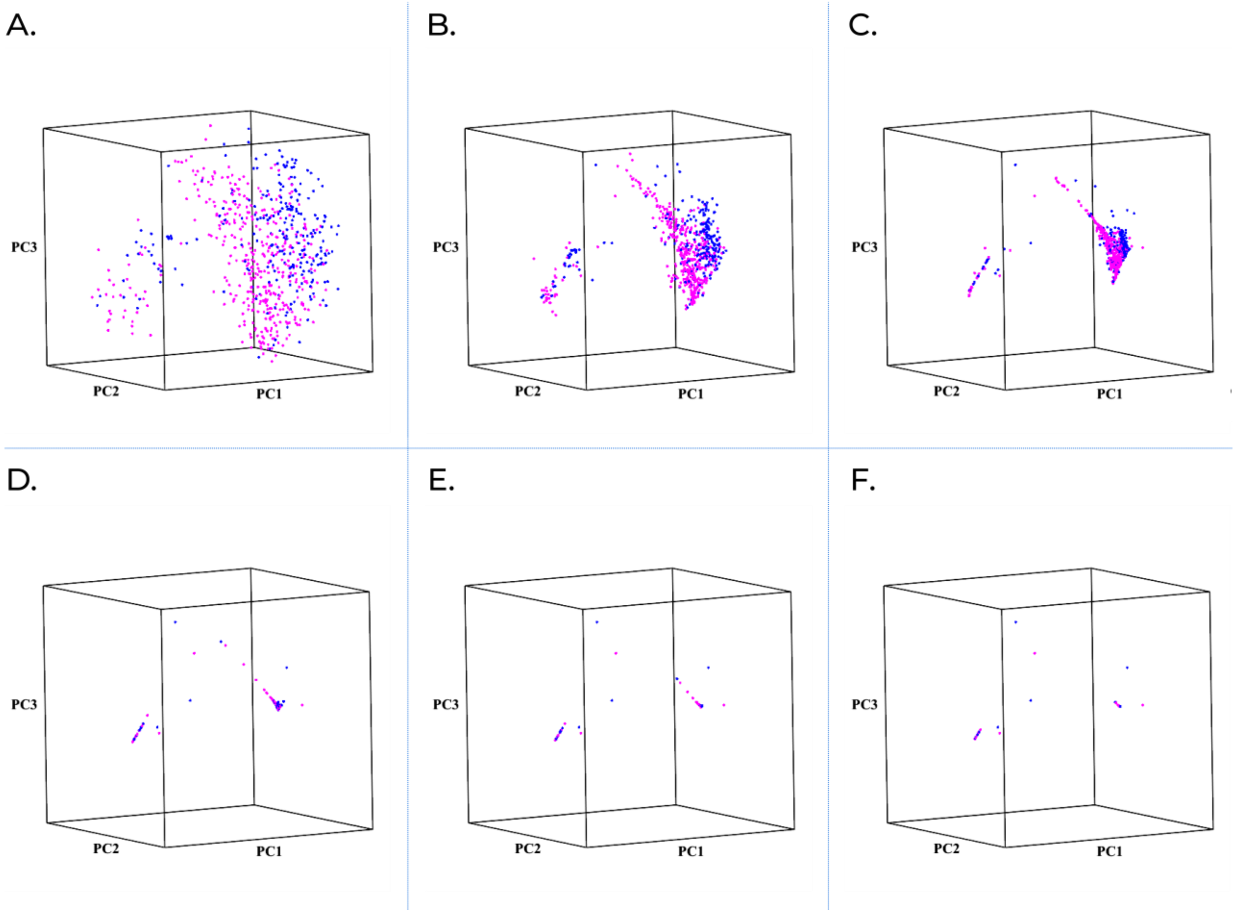


**Supplementary Figure S8. Direct analysis of shared TCGA–CGGA genes does not clearly separate CGGA gliomas.**

DQC was applied directly to the 657 CGGA tumors with available overall-survival data using the 16,154 genes shared between TCGA and CGGA. Each point represents one tumor. Tumors are shown in a three-dimensional view for display only. Clinical information, including histology, recurrence status, and survival, was not used to build the tumor map. Colors indicate histology added afterward for interpretation: magenta, LGG; blue, GBM. **(A–F)** Six representative frames show how tumors moved during DQC analysis. Although tumors became more tightly grouped over time, LGG and GBM tumors remained mixed rather than separating into clear clinical groups. These results show that direct analysis of the shared gene set did not recover a clinically useful glioma organization in CGGA, supporting the need for the BioCoord-based validation analysis.

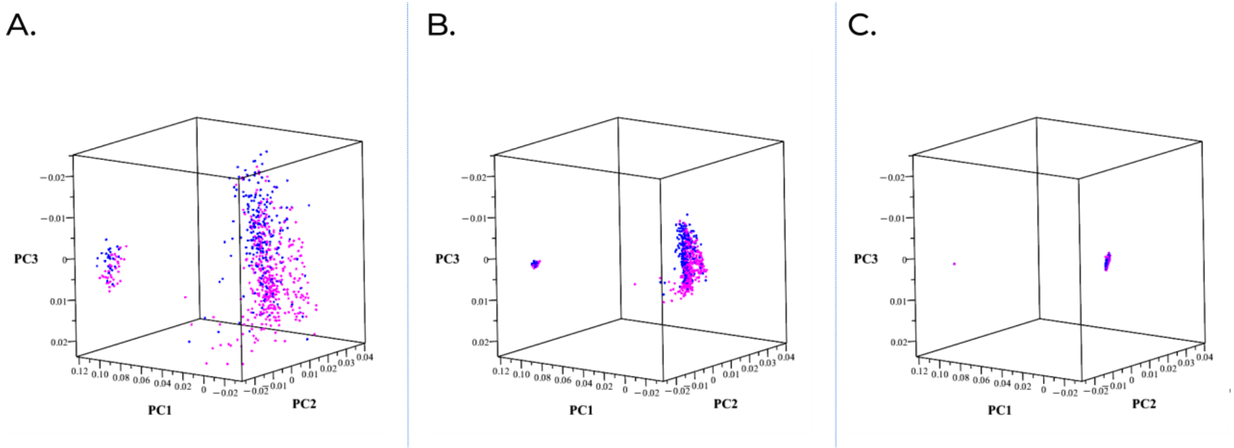


**Fig. S9. Denoising CGGA gene-expression data by feature selection improves visual clustering but does not recover diagnostic or prognostic structure.**

To test whether reducing noise in the CGGA gene-expression matrix could improve clustering performance, DQC was applied to a knee-selected reduced gene set of approximately 636 genes. Representative DQC evolution frames are shown: (A) initial frame, (B) intermediate frame, and (C) late frame. Each point represents one tumor. For visualization only, diagnostic labels were applied post hoc: GBM samples are shown in blue, and LGG samples are shown in magenta. These diagnostic annotations were not used for clustering. Compared with the full shared gene-expression matrix, the reduced gene set produced clearer visual contraction and limited local organization during DQC evolution. However, unlike the TCGA discovery analysis, this feature-selected CGGA gene-expression map did not resolve a clear diagnostic structure: LGG and GBM samples remained substantially overlapping, and the resulting clusters did not yield meaningful overall-survival stratification in post hoc analyses. These findings show that feature selection can reduce visual noise, but gene-expression clustering alone was not sufficient to recover clinically meaningful glioma organization in the independent CGGA cohort.

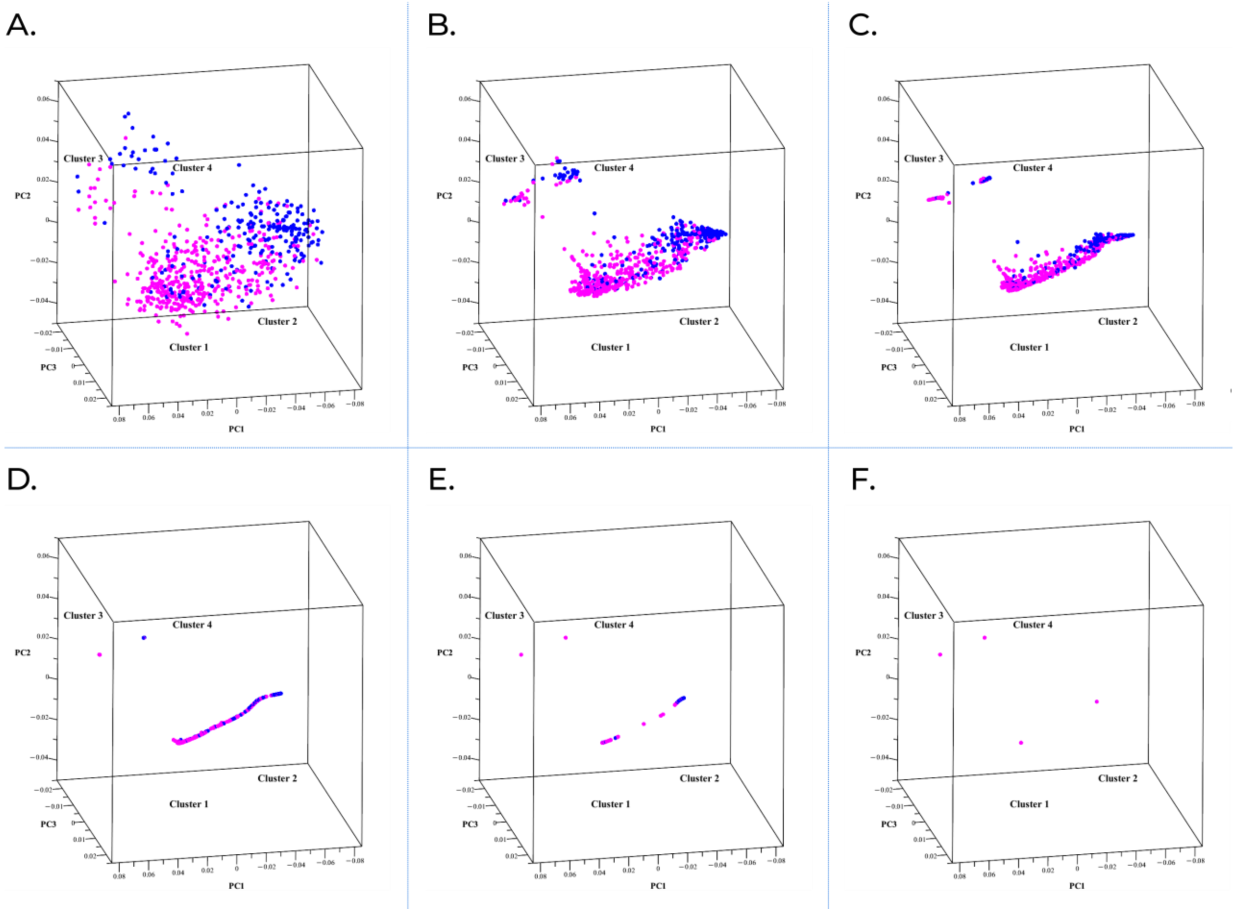


**Supplementary Fig. S10. CGGA tumors organize into clinically recognizable groups in TCGA-derived BioCoord space.**

DQC was applied to 693 CGGA tumors using four BioCoords recalculated from CGGA RNA expression with the TCGA-defined BioCoord gene modules. For visualization only, tumors are displayed in the first three principal components of the four-dimensional BioCoord space. Representative DQC frames are shown: (A) initial frame, (B) early frame, (C) intermediate frame, (D–E) late frames, and (F) final frame. Each point represents one tumor. Diagnostic labels, survival, and other clinical annotations were not used during clustering. For post hoc visualization, diagnostic labels were overlaid after clustering: LGG samples are shown in magenta and GBM samples in blue. Cluster labels indicate the approximate locations of the final DQC-defined tumor groups. During DQC evolution, CGGA tumors move from a dispersed pattern into a more organized BioCoord-based tumor map, with two major tumor populations and two smaller separated groups. Intermediate frames best show the relationship among tumor states, whereas later frames show contraction toward final group positions. This analysis shows that TCGA-derived BioCoords preserve clinically recognizable glioma organization in an independent CGGA cohort.

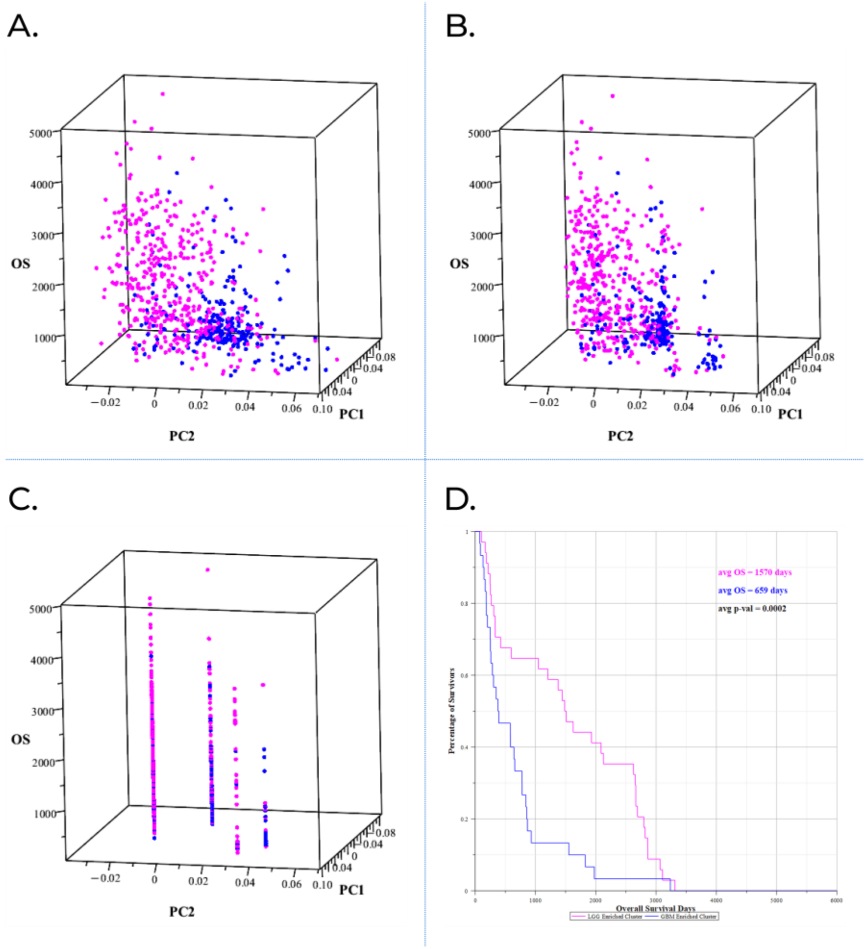


**Supplementary Fig. S11. Overall survival aligns with BioCoord-derived tumor organization in CGGA.**

CGGA tumors were projected into TCGA-derived BioCoord space, and overall survival was overlaid after DQC-based tumor groups were defined. Clinical outcome was not used during BioCoord calculation or clustering. (A–C) Three-dimensional visualizations show overall survival distribution across the BioCoord-derived tumor map from different viewing angles. Each point represents one tumor; the vertical axis shows overall survival time. Diagnostic labels are shown post hoc, with LGG samples in magenta and GBM samples in blue. These views show that tumors with different survival outcomes occupy different regions of the BioCoord map, rather than being randomly distributed across the space. (D) Kaplan–Meier analysis comparing BioCoord-derived tumor groups shows clear overall-survival separation, with longer survival in the LGG-enriched BioCoord group and shorter survival in the GBM-enriched BioCoord group. Together, these results show that BioCoord-derived tumor organization in CGGA is associated with overall survival.

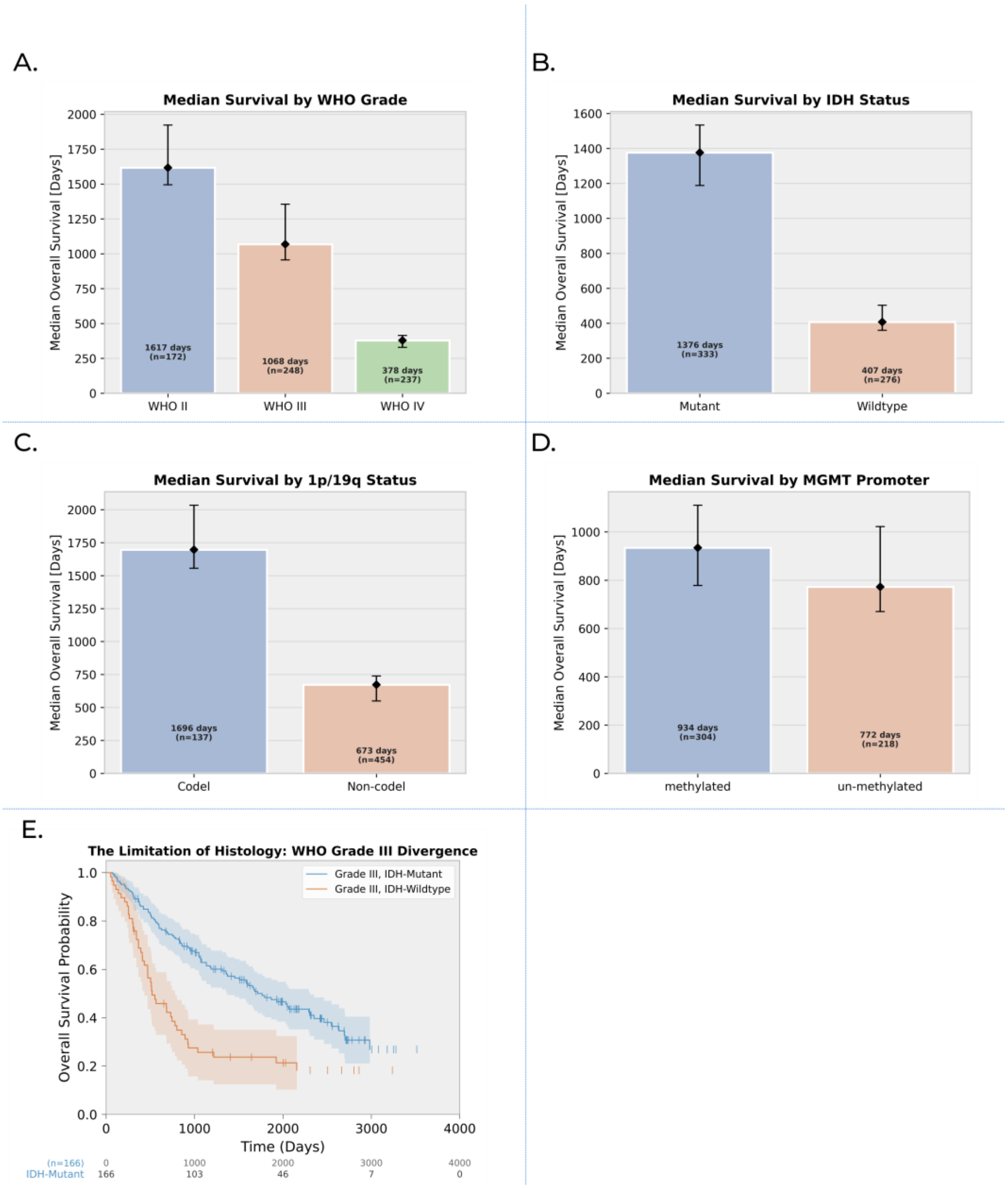


**Supplementary Fig. S12. Established clinical and molecular features are associated with overall survival in CGGA but do not fully capture outcome heterogeneity.**

To place the BioCoord findings in the context of standard clinical and molecular predictors, overall survival was examined across key CGGA annotations. (A) Median overall survival stratified by WHO grade shows progressively shorter survival from Grade II to Grade IV tumors. (B) Median overall survival stratified by IDH mutation status shows longer survival in IDH-mutant tumors compared with IDH-wildtype tumors. (C) Median overall survival stratified by 1p/19q status shows longer survival in codeleted tumors compared with non-codeleted tumors. (D) Median overall survival stratified by MGMT promoter methylation status shows a modest survival difference between methylated and unmethylated tumors. (E) Kaplan–Meier analysis restricted to WHO Grade III tumors shows clear survival divergence by IDH status, indicating that tumors within the same histologic grade can have different clinical outcomes. Together, these analyses show that established diagnostic and molecular variables are clinically informative, while also highlighting survival heterogeneity that is not fully captured by any single variable alone.

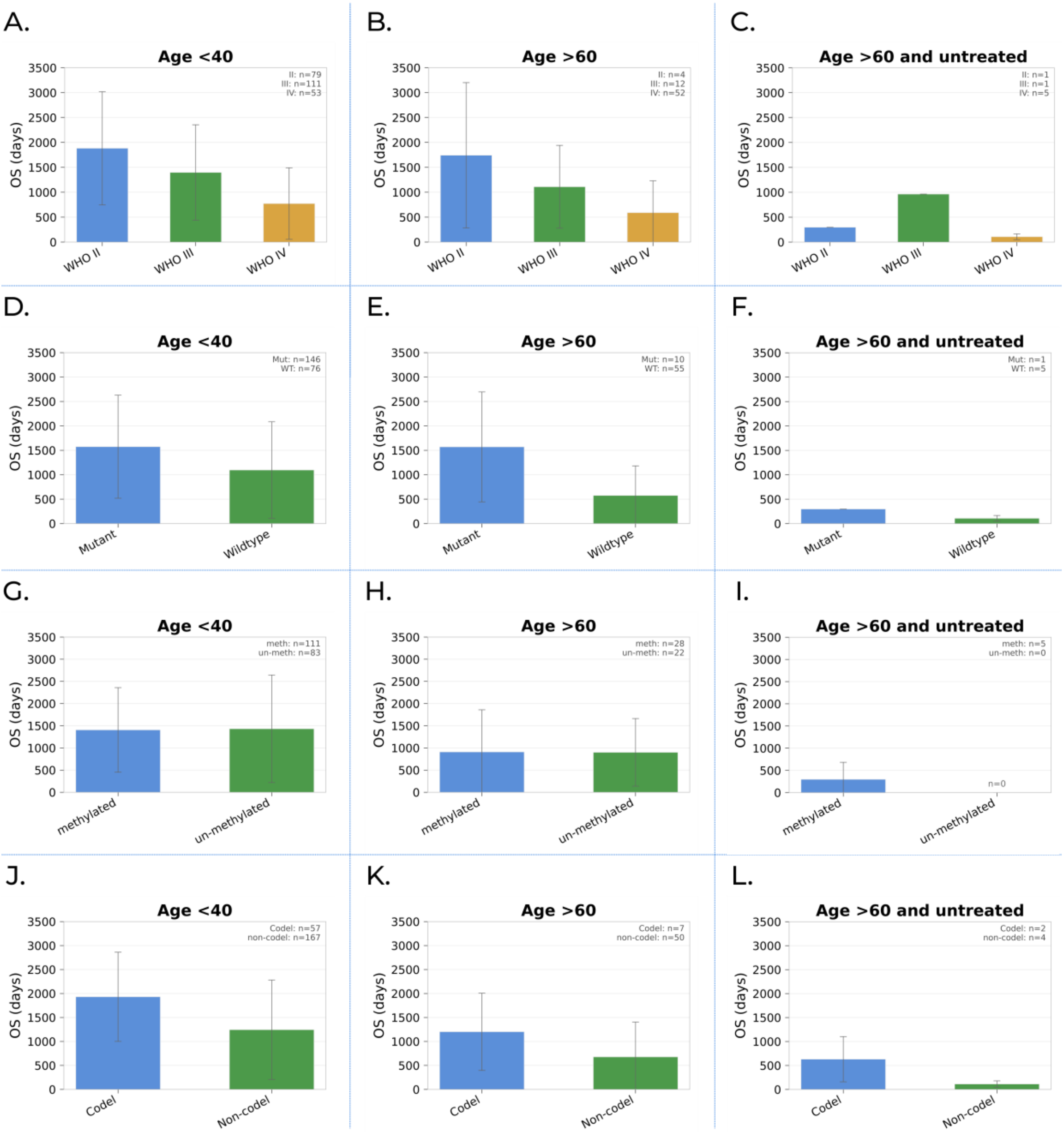

**Supplementary Fig. S13. Survival effects of clinical and molecular features vary across age and treatment contexts in the CGGA cohort.**

To assess whether established prognostic features show consistent survival patterns across clinical contexts, median overall survival (OS, days) was examined after stratification by age and treatment status. **(A–C)** Median OS by WHO grade is shown for patients aged <40 years, patients aged ≥60 years, and patients aged ≥60 years without treatment, respectively. **(D–F)** Median OS by IDH mutation status is shown across the same three patient subsets. **(G–I)** Median OS by MGMT promoter methylation status is shown across the same three patient subsets. **(J–L)** Median OS by 1p/19q codeletion status is shown across the same three patient subsets. Across these analyses, expected survival trends were generally observed, including shorter survival with higher WHO grade and longer survival with IDH mutation or 1p/19q codeletion. However, the magnitude of these survival differences varied by age and treatment context, indicating that no single clinical or molecular feature fully accounts for outcome heterogeneity in CGGA.
