## Supplementary material for "Dynamic Quantum Clustering of Glioma RNA-seq Reveals Interpretable Tumor States Linked to Diagnosis and Survival": Table S5

**Supplementary Table S5. DQC cluster membership and survival summaries for top-k gene panels**

Notes: Clusters are derived by DQC without labels. TCGA histology and PFS/OS were applied post hoc for evaluation only. PFS/OS values are reported as mean days.

**Table S5-1-1. TCGA histology within the three groups identified by DQC using 140 genes**

|  | **Cluster 140A** | **Cluster 140B** | **Cluster 140C** |
| --- | --- | --- | --- |
| TCGA LGG | 450 | 9 | 65 |
| TCGA GBM | 0 | 168 | 0 |

**Table S5-1-2. Mean progression-free and overall survival in the 140-gene DQC groups**

| **Cluster** | **Mean PFS, days** | **Mean OS, days** |
| --- | --- | --- |
| Cluster 140A | 811.01 | 1053.23 |
| Cluster 140B | 275.26 | 444.76 |
| Cluster 140C | 389.89 | 659.86 |

**Table S5-2-1. TCGA histology within the four groups identified by DQC using 100 genes**

|  | **Cluster 100A** | **Cluster 100B** | **Cluster 100C** | **Cluster 100D** |
| --- | --- | --- | --- | --- |
| TCGA LGG | 421 | 5 | 65 | 33 |
| TCGA GBM | 0 | 168 | 0 | 0 |

**Table S5-2-2. Mean progression-free and overall survival in the 100-gene DQC groups**

| **Cluster** | **Mean PFS, days** | **Mean OS, days** |
| --- | --- | --- |
| Cluster 100A | 835.49 | 1061.96 |
| Cluster 100B | 262.65 | 430.05 |
| Cluster 100C | 397.34 | 602.26 |
| Cluster 100D | 448.42 | 1043.55 |

**Table S5-3-1. TCGA histology within the four groups identified by DQC using 90 genes**

|  | **Cluster 90A** | **Cluster 90B** | **Cluster 90C** | **Cluster 90D** |
| --- | --- | --- | --- | --- |
| TCGA LGG | 421 | 5 | 65 | 33 |
| TCGA GBM | 0 | 168 | 0 | 0 |

**Table S5-3-2. Mean progression-free and overall survival in the 90-gene DQC groups**

| **Cluster** | **Mean PFS, days** | **Mean OS, days** |
| --- | --- | --- |
| Cluster 90A | 836.31 | 1062.83 |
| Cluster 90B | 262.65 | 430.05 |
| Cluster 90C | 405.85 | 691.82 |
| Cluster 90D | 457.94 | 871.12 |

### ****Table S5. DQC cluster composition by TCGA histology and mean survival for the 140-, 100-, and 90-gene panels****

Genes from the 554-gene panel were ranked by two-sample t-test P values between the two DQC-defined groups. The test was used only to order the genes. DQC was then repeated using the top 140, 100, and 90 genes. TCGA histology and survival information were not used to form the clusters but were added afterward to evaluate their clinical relevance.

Using 140 genes, DQC identified three clusters: two LGG-only groups with different mean survival and one GBM-enriched group containing all 168 GBMs and nine LGGs. Using 100 genes, DQC identified four clusters: three LGG-only groups and one GBM-enriched group containing all 168 GBMs and five LGGs. The 90-gene panel preserved the same four-cluster histologic composition and the survival order A → D → C → B.

Together, these results show that reducing the gene set to 90 genes preserved the main LGG–GBM separation while revealing LGG groups with distinct clinical outcomes. Among the panels presented in this table, the 90-gene panel was the smallest that preserved this four-group pattern. When fewer than 90 genes were used, the four-cluster pattern was no longer consistently preserved (data not shown).
